# Novel blood-based proteomic signatures across multiple neurodegenerative diseases

**DOI:** 10.1101/2024.12.28.24319680

**Authors:** Robert Durcan, Amanda Heslegrave, Peter Swann, Julia Goddard, Leonidas Chouliaras, Alexander G Murley, George Savulich, W Richard Bevan-Jones, Owen Swann, Nicholas J Ashton, Kaj Blennow, William McEwan, Henrik Zetterberg, James B Rowe, John T O’Brien, Maura Malpetti

## Abstract

**INTRODUCTION:** Blood-based biomarkers have the potential to support early and accurate diagnosis of neurodegenerative diseases, which is sensitive to molecular pathology and predictive of outcome. We evaluated a novel multiplex proteomic method in people with diverse neurodegenerative diseases.

**METHODS:** Serum from people with Alzheimer’s disease (N=36), Lewy body dementia (N=34), frontotemporal dementia (N=36) and progressive supranuclear palsy (N=36) and age-matched controls (N=30) was analysed with the NUcleic acid Linked Immuno-Sandwich Assay (NULISA) central nervous system panel (∼120 analytes) and inflammation panel (250 analytes). Biomarkers were compared across groups and included as predictors of survival.

**RESULTS:** The NULISA panels demonstrated high sensitivity and reliability for detecting multiple biomarkers across neurodegenerative disorders. There were condition-specific proteomic biomarkers, while NfL, CRH, CD276 and S100A12 were significant transdiagnostic outcome predictors.

**DISCUSSION:** The sensitive NULISA multiplex approach supports differential diagnosis and target identification, with prognostically informative dementia-related biomarkers.

## 1. Background

As the field of neurodegenerative dementias and movement disorders moves into an era of disease modifying therapies, there is increasing need for provide early and accurate diagnoses. Current clinically used Alzheimer’s disease (AD) biomarkers include cerebrospinal fluid (CSF) measures of Aβ40/42 and phosphorylated tau, and amyloid- and tau-PET scans [1,2]. Both CSF and PET markers have high cost, infrastructure, and patient-tolerance barriers to implementation at scale in “real-world” settings. Seed amplification assays (SAA) using biofluids and skin biopsies are highly specific to distinct synucleinopathies[3] and tauopathies [4], but issues of co-pathology and scalability remain challenging.

Blood-based proteomic biomarkers offer a promising avenue for early and accurate diagnosis in neurodegenerative diseases that is scalable and deliverable in routine clinical care. Blood tests are very well tolerated, can be performed repeatedly for an individual, allowing assessment of test-retest reliability and temporal profiles through longitudinal assessment of disease trajectory, or in response to treatment.

Blood-based proteomic biomarkers have the potential to support diagnoses based on specific molecular pathologies of disease; or concomitant pathophysiological processes such as neuroinflammation and synaptic loss. The clinical and pathological heterogeneity seen within and between diagnostic groups may also be reflected in biomarker signals. Disease-specific fingerprints or disease-agnostic pathways may provide insights into mechanisms of disease and potential therapeutic targets. The revolution of blood-based biomarkers in dementia has been rapid in the last 5 years [5–10], despite the low concentration of many proteins in circulating blood; and the limitation inherent in quantification of one marker or a small set of markers a time.

Recently, proteomic methods have been developed with high-sensitivity multiplex approaches. For example, the <u>Nu</u>cleic acid-<u>L</u>inked Immuno-<u>S</u>andwich <u>A</u>ssay (NULISA), is a novel method of measuring many molecules in the human plasma and serum proteome, even to attomolar concentrations [11]. The NULISAseq “CNS Disease panel” analyses over 120 proteins associated with diverse neurodegenerative diseases. Results from this technique have been reported in Alzheimer’s Disease (AD) [12–14]. The NULISAseq “Inflammation Panel” analyses 250 proteins, many of which have been associated with the inflammatory responses observed neurodegenerative diseases [11].

We took a transdiagnostic approach to assess the utility of the NULISAseq method, with two principal aims. First, we compared NULISAseq assays of conventional biomarkers of neurodegeneration (phosphorylated tau isoforms, neurofilament light chain (NfL / NEFL) and glial fibrillary acidic protein (GFAP)) against results from the Quanterix Single molecule array (SIMOA) [7–10] with blood samples collected simultaneously from the same participants. Second, we integrated results from both NULISAseq CNS and Inflammatory panels to identify differentially expressed diagnostic and prognostic markers in people with well-defined common neurodegenerative diseases, including AD, Lewy body dementia (LBD), frontotemporal dementia (FTD) and progressive supranuclear palsy (PSP).

## 2. Methods

### 2.1. Participants

Patient participants were recruited from specialist clinics for cognitive and movement disorders at the Cambridge University Hospitals NHS Trust and collaborating regional psychiatry and neurology services. We included participants with a clinical diagnosis that met clinical consensus diagnostic criteria for MCI (n=20, amyloid biomarker positive) or AD-dementia (n=16) [15,16], probable or possible PSP (n=36 ) [17], FTD (n=36; 21 with behavioural variant FTD and 15 with primary progressive aphasia (PPA), consisting of 8 with non-fluent variant PPA and 7 with semantic variant PPA) [18,19], and LBD (n=34; 29 with dementia with Lewy body and 5 with Parkinson’s disease dementia) [20]. As with previous studies from this patient participant cohort [21–23], we considered patients with AD and Aβ-positive MCI as a single cohort (AD/MCI+) as they represent a biological continuum. In a sub-group of participants (n=48, 23 AD/MCI+, 25 LBD), PET and/or CSF markers for amyloid were available to confirm the presence or absence of β-amyloid (interpreted as AD-pathology) in conditions with weak clinicopathological correlations and either high likelihood of a significant fraction with AD as main or co-pathology (LBD) or a high likelihood of clinical diagnostic false positives (amnestic MCI). Of the MCI patients included in the study, all were amyloid-positive with PET imaging with the Pittsburgh compound B (PiB tracer at a cut-off of 19 centiloids [24]) and/or CSF Alzheimer’s biomarkers at lumbar puncture (Amyloid-beta 1-42/40 ratio < 0.065 as recommended laboratory threshold from University College London Hospitals reference laboratory [25]). These participants were analysed together with those with early AD dementia as a single AD group (AD/MCI+). Of the LBD cohort, n=17/25patients with PiB PET were amyloid-positive. We also included n=30 healthy controls with MMSE>26/30, absence of memory symptoms and no significant medical illnesses[26]. Exclusion criteria for both patient and healthy control cohorts included recent or current acute infection, major concurrent psychiatric illness, other severe physical illness, or a history of other significant neurological illness and/or autoimmune conditions. Participants underwent baseline clinical and neuropsychological assessment, including the revised Addenbrooke’s Cognitive Examination (ACE-R, 0-100 points). Survival data were collected up to census date of 24^th^ of November 2024.

### 2.2. Ethical approval and consent to participate

Participants with mental capacity gave their written informed consent to take part in the study according to the Declaration of Helsinki. For those who lacked capacity, their participation followed the personal consultee process in accordance with the UK law. The research protocols were approved by the National Research Ethics Service’s East of England Cambridge Central Committee (REC references: 07/Q0102/3; 13/EE/0104).

### 2.3. Blood sample collection and processing

Blood samples were obtained by venepuncture and collected in EDTA tubes. They were centrifuged to isolate plasma and serum, aliquoted and stored at −70/80 °C until further analyses.

Serum samples from all patients and controls were analysed with NULISA. The CNS panel (see Supplementary Table 1 for panel list), including Aβ peptides, phosphorylated tau forms, NfL (or ‘NEFL’ as from the panel nomenclature) and other markers of neurodegeneration, inflammation, and vascular health. In addition to the CNS panel, the samples were also analysed with the NULISAseq Inflammation Panel 250 (full list in Supplementary Table 2) for simultaneous analysis of 250 proteins associated with inflammatory and immune response processes (e.g. interleukins, chemokines, complement) with attomolar sensitivity to fg/ml units[11]. Sample and data analysis was performed according to kit manufacturer protocols, including Log2 Transformation of the data and NULISA Protein Quantification (NPQ) on the logarithmic scale (Alamar Biosciences, Fremont, CA).

The majority (91%) of participants (n=156; Control=28, AD/MCI+=34 (MCI=19, AD=15), FTD=32 (bvFTD=18, PPA=14), LBD=30, PSP=32), also had paired plasma samples, that were analysed at the Clinical Neurochemistry Laboratory in Mölndal (Sweden). Plasma samples were thawed on wet ice, centrifuged at 500×□g for 5□min at 4°C. Calibrators (neat) and samples (plasma: 1:4 dilution) were measured in duplicates. The plasma assays performed were the Quanterix Simoa Human Neurology 4-Plex E assay, measuring Aβ40, Aβ42, GFAP and NfL (Quanterix, Billerica, MA), p-tau231 and the p-tau217 ALZpath assay, as previously described [5]. Plasma samples were analyzed at the same time using the same batch of reagents. A four-parameter logistic curve fit data reduction method was used to generate a calibration curve. Two control samples of known concentration of the protein of interest (high-control and low-control) were included as quality control. Intra-assay coefficients of variation were below 10%.

### 2.4. Statistical analysis

Analyses were performed using R (version 4.4.2). For descriptive statistics chi-square and analysis of variance tests compared variables between groups. Statistical analyses on blood-derived markers were carried out in three steps.

First, we compared NULISA CNS panel results on serum samples with Simoa assays on plasma samples from the same participants. Since NULISA NPQ values are log2-transformed, we applied log2 transformation to the absolute quantification values of plasma markers (as log2(pg/mL + 1)). Following transformation, we performed Spearman’s correlation analyses on like-for-like markers, including p-tau217, p-tau231, NfL and GFAP. Then, we ran group comparisons with Kruskal-Wallis tests, with Dunn’s post-hoc tests, on NULISA single markers separately.

Second, we implemented Linear Models for Microarray and RNA-Seq Data (LIMMA models) to compare the differential protein expression on each panel, between diagnostic groups and controls.. Results were displayed with volcano plots using colour schemes to highlight the markers that were significantly different between groups, considering uncorrected p values and after false discovery rate (FDR) correction for multiple comparisons. All LIMMA analyses included age and sex as covariates.

Third, we investigated the relationship between dementia-relevant markers on the CNS panel and survival. Survival status was available for all the patient participants who passed quality check for the CNS panel (n=141, 70 deceased and 71 still alive) at the census date. Survival analysis used Cox proportional hazards regression (R function coxph), including levels of NfL, p-tau217, p-tau231, GFAP, and years from blood test to death as moderating variables of interest, and age, sex and diagnosis group as covariates. We then applied a similar analysis with the markers from the inflammation panel (n=138 patients: 68 deceased and 70 still alive) that were significantly increased across all patients as compared to controls.

### 2.5. Data Availability

Anonymized processed data can be shared upon request to the corresponding author. Raw data may also be requested but are likely to be subject to a data transfer agreement with restrictions required to comply with participant consent and data protection regulations.

## 3. Results

### 3.1. Cohort characteristics

Participant demographics and clinical characteristics are described in Table 1. Patients with LBD were slightly older than controls and had the highest proportion of male participants. Participants with AD/MCI+ and PSP scored significantly higher on the ACE-R than other groups, while patients with LBD and FTD showed the lowest performance on average.

**Table 1.** Demographic and clinical characteristics for control and patient groups (* = significant difference between patient group and controls in post hoc analysis).

| Group | N | Age | Sex (F/M) | ACE-R (score/100) |
| --- | --- | --- | --- | --- |
| Control | 30 | 68.3 (6.1) | 14/16 | - |
| AD/MCI+ | 36 | 71.8 (8.1) | 16/20 | 79.0 (10.7) |
| LBD | 34 | 73.9 (6.9)* | 6/28 | 69.4 (14.8) |
| FTD | 36 | 64.5 (7.8) | 12/24 | 67.7 (20.4) |
| PSP | 36 | 69.4 (6.7) | 17/19 | 83.4 (10.2) |
| Group comparisons | - | F(4)=8.55, p<0.001 | X<sup>2</sup>(4)=9.22, p=0.06 | F(3)=9.37, p<0.001 |

### 3.2. Quality control checks

NULISA markers were detected with high sensitivity in both CNS and Inflammation panels (94.1% and 97.3% target detectability, respectively). Two samples (1 control, 1 PSP) failed the quality check for the CNS panel and four samples (1 DLB, 1 nfvPPA, 2 PSP) for the Inflammation panel. These were excluded from further analyses. On the main CNS markers (p-tau217, p-tau231, NfL and GFAP) we tested for associations with freezer storage time, as data collection spanned 10 years. Correlation analyses between NULISA CNS protein quantification and years of storage in –70/-80 °C freezers did not identify significant associations (See Supplementary Figure 1).

### 3.3. NULISA vs Simoa biomarker measurements

We compared NULISA serum-based markers with Simoa plasma-based markers. All principal like-for-like proteins significantly correlated (Figure 1A). Specifically, p-tau217 and NfL showed the strongest correlations (rho=0.798 and rho=0.881 respectively, p<0.0001), followed by GFAP (rho=0.762, p<0.0001) and p-tau231 (rho=0.693, p<0.0001).

**Figure 1.**
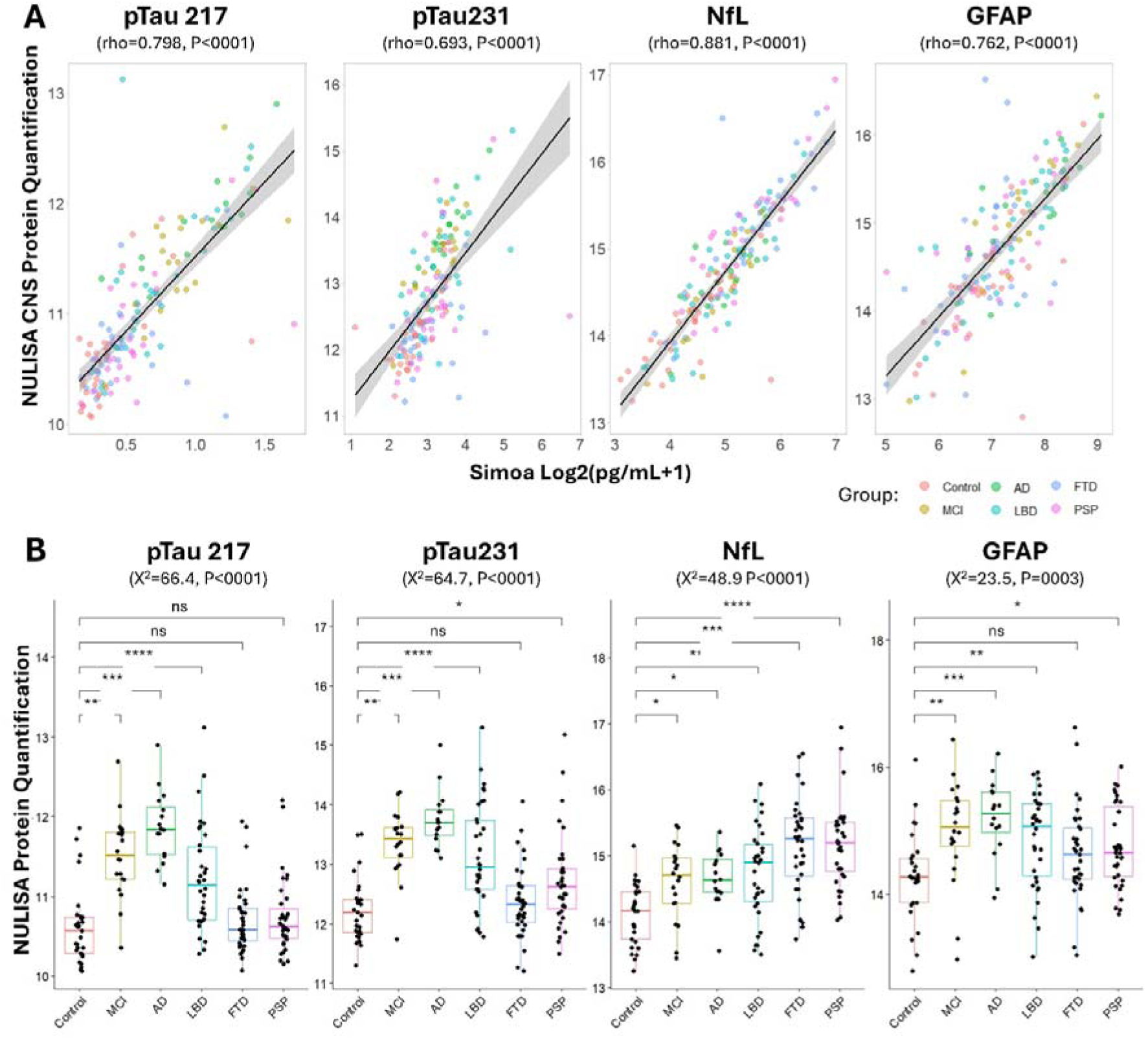
Validation of the dementia-relevant single markers on the CNS panel: A) Associations between NULISA serum-based markers and Simoa plasma-based markers, considering like-for-like proteins. B) Group comparisons on dementia-relevant single markers from the NULISA CNS panel.

### 3.4. NULISA CNS panel across diagnostic groups

Group comparisons on dementia-relevant single markers from the NULISA CNS panel revealed diagnostic differences (Figure 1B). Specifically, Kruskal Wallis Tests were significant across all 4 markers. Post hoc Dunn tests (considering adjusted p values) indicated that p-tau217 levels were significantly increased in patients with AD/MCI+ and LBD as compared to controls, while p-tau231 levels were slightly increased also in patients with PSP. Levels of NfL were higher in all patient groups as compared to controls, while GFAP levels were increased especially in patients with AD/MCI+ and LBD and to a lower extent in patients with PSP.

Comparing each patient cohort with controls (Figure 2) including the whole CNS panel, p-tau epitopes (p-tau217, p-tau231, p-tau181) were significantly increased in AD/MCI+ and LBD. In patients with AD/MCI+, but not LBD, serum levels of MAPT were also significantly increased as compared to controls. Increased levels of serum NfL was the most significant marker in comparing FTD and PSP cohorts with controls, followed by multiple cytokines and chemokines. The homeostatic autophagy regulating scaffold protein (SQSTM1) was elevated in PSP versus controls and AD/MCI+, while Presenilin 1 (PSEN1), a key protein in amyloid-β clearance, was elevated in both FTD and PSP cohorts versus controls. PARK7 and PGK1 levels, important enzymes in glycolysis regulation, were increased in FTD versus controls. All patient groups showed lower levels of corticotropin-releasing hormone (CRH), than controls. This survived FDR correction in the comparison between patients with FTD and controls. See Supplementary Table 3 for all comparisons.

**Figure 2.**
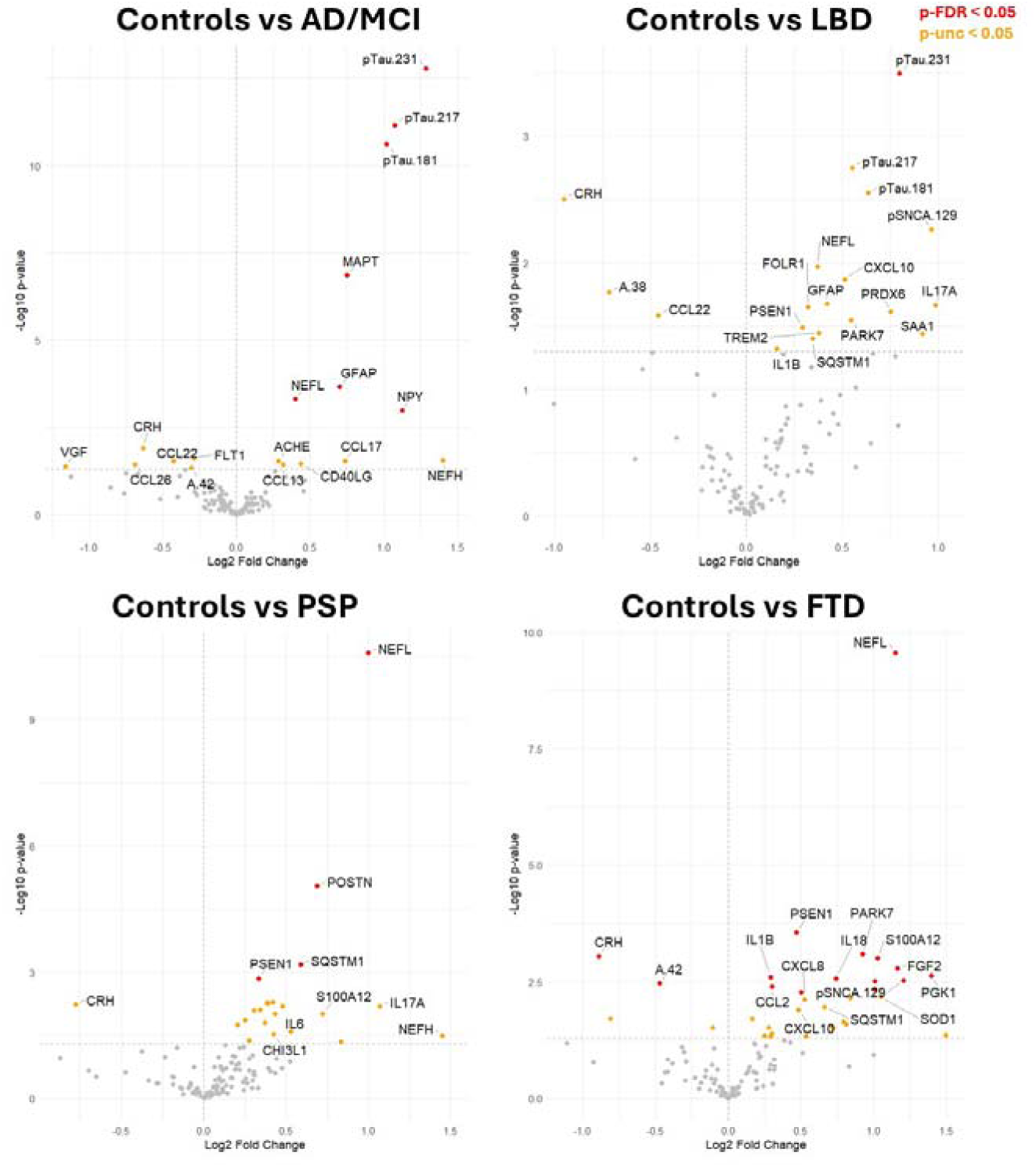
Comparisons between each patient group and controls on CNS markers. For each comparison, the panel on the right represents markers that are increased in patients as compared to controls, while the panel on the left represents markers reduced in patients. Orange dots represent comparisons with p<0.05, and red dots with p<0.05 corrected for multiple comparisons.

Next, the diagnostic groups were analysed against the AD/MCI+ group (Supplementary Figure 2). AD/MCI+ had significant higher levels of p-tau217, p-tau231, p-tau181, MAPT and pro-neuropeptide Y (NPY) as compared to FTD and PSP cohorts. Comparing FTD and PSP, patients with PSP showed higher levels of inflammation-related markers, including IL7, CX3CL1, and periostin (POSTN) than FTD. In FTD, higher levels of fibroblast growth factor 2 (FGF2) were observed than in PSP. Compared to the AD/MCI+ cohort (Supplementary Figure 2), patients with LBD showed higher levels of neuropentraxins (NPTX1/2) than AD/MCI+, but these comparisons did not survive FDR correction.

### 3.5. NULISA Inflammation panel across diagnostic groups

Comparing each patient cohort with controls (Figure 3), increased levels of Amphiregulin (AREG) and CD276 (also known as B7-H3) were identified in LBD, FTD and PSP as compared to controls. Patients with FTD and PSP showed higher levels in multiple inflammation markers, while AD/MCI+ and LBD cohorts showed a more limited pattern of upregulated markers. Patients with AD/MCI+ showed increased GFAP levels, but this comparison did not survive FDR correction. Importantly, C-reactive protein levels, a commonly used clinical marker of non-specific systemic inflammation, did not show any significant differences between diagnostic groups and controls. See Supplementary Table 4 for all comparisons.

**Figure 3.**
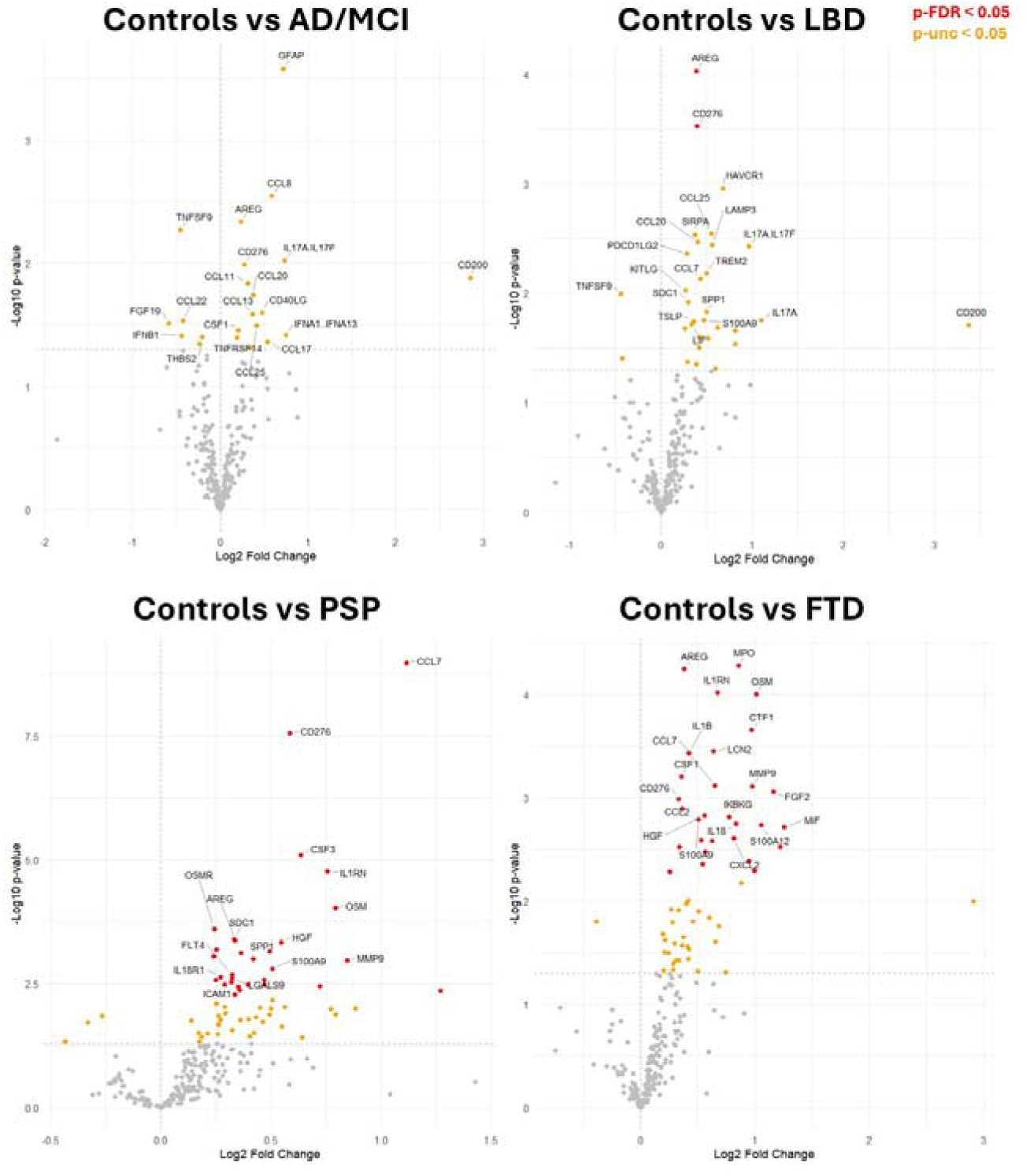
Comparisons between each patient group and controls on inflammation markers. For each comparison, the panel on the right represents markers that are increased in patients as compared to controls, while the panel on the left represents markers reduced in patients. Orange dots represent comparisons with p<0.05, and red dots with p<0.05 corrected for multiple comparisons.

Comparing each patient group to the AD/MCI+ cohort (Supplementary Figure 3), patients with LBD showed similar levels of inflammation markers as AD/MCI+, with a larger number of markers being upregulated in LBD than AD/MCI+ when FDR correction was not considered. FTD and PSP cohorts had higher levels in several inflammation markers as compared to AD/MCI+, including Matrix metalloproteinase-9 (MMP9) and Hepatocyte Growth Factor (HGF). Comparing FTD and PSP, no marker was significantly different when FDR correction was considered.

### 3.6. NULISA markers for predicting survival

We tested dementia-relevant markers from the CNS panel as prognostic biomarkers (in patients only) using Cox proportional hazards regression with NfL, p-tau217, p-tau231, GFAP and time in days from blood test to death as predictors of interest for survival (deceased/alive), and age, sex and disease groups as covariates (Figure 4A). NfL levels were significantly associated with shorter time to death [hazard ratio 2.09 (1.35–3.23), p=0.001]. As lower CRH levels were found in all patient groups as compared to controls, we also tested their association with survival rates, correcting for NfL, age, sex and diagnosis. CRH levels were inversely associated with time to death [hazard ratio 0.76 (0.60-0.96), p=0.022]. To illustrate the prognostic effect of NfL (Figure 4A) and CRH (Figure 4B), we plotted patients with high and low values of NfL and CRH, as separated by the median. These effects were also plotted by disease groups in Supplementary Figure 4.

**Figure 4.**
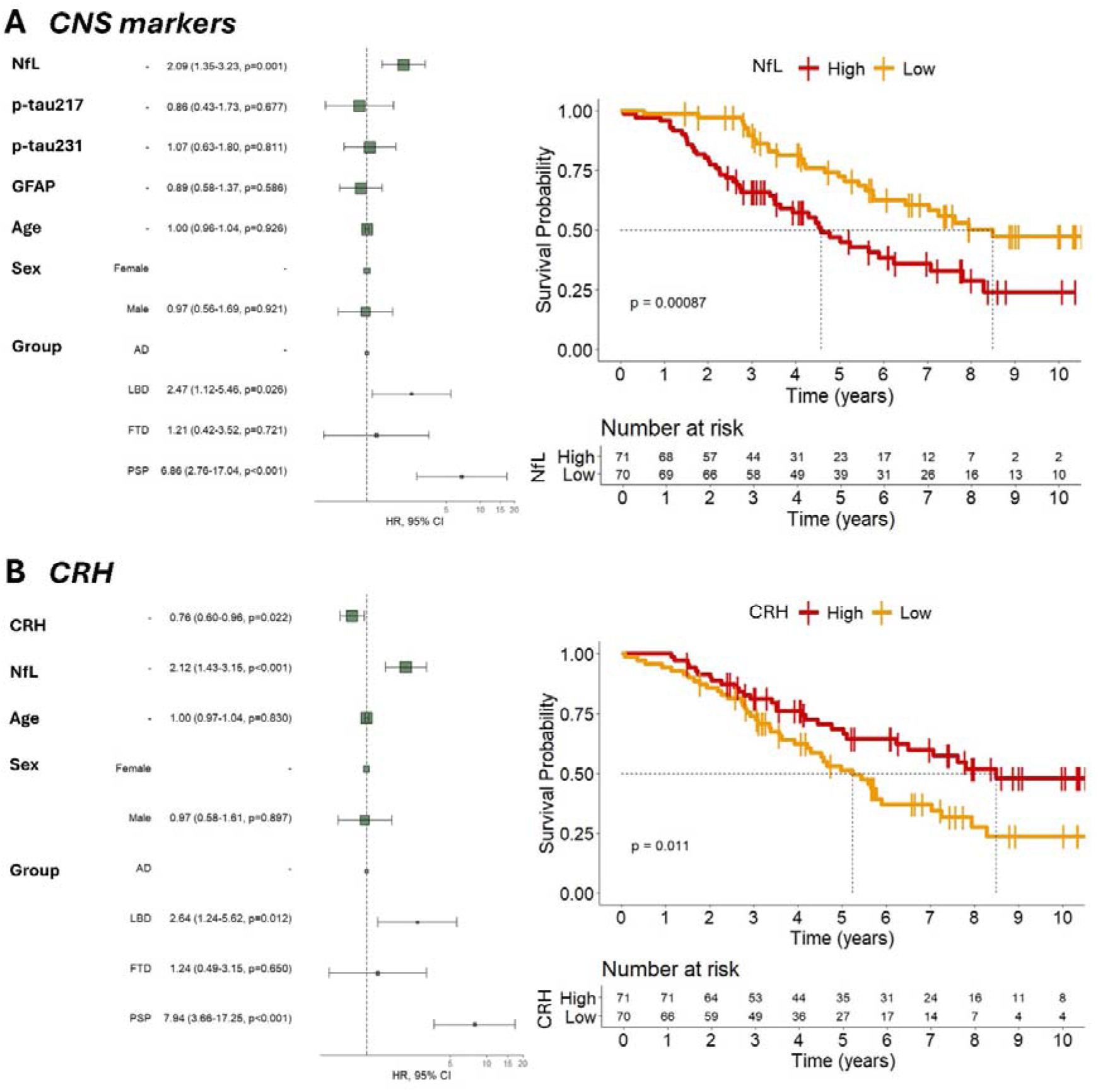
Prognostic markers on CNS panels. A) Survival analysis including the main dementia-related markers, age and diagnosis as predictors, and Kaplan–Meier Survival Curve of NfL divided on the median; B) Survival analysis including the most significant inflammation markers, age and diagnosis as predictors, and Kaplan–Meir Survival Curve of CRH divided on the median.

We identified 13 markers from the inflammation panel that were significantly increased in patients (across all groups) as compared to controls, considering FDR-corrected p values (see Supplementary Figure 5): AREG, CD276, CCL7, GFAP, IL1RN, CSF1, OSM, CSF3, CTF1, IL17A-IL17F, S100A12, and TSLP. In contrast, TNFSF9 was decreased in all patient groups. We included these markers as predictors of interest for survival alongside days from blood test to death in a Cox proportional hazards regression, and age, sex and disease groups were included as covariates. Given the high number of predictors, we fitted the full model and performed backward stepwise selection, which compared the full model with reduced models based on the Akaike information criterion (AIC) until no further improvement in AIC is achieved excluding predictors. The model selection retained CD276, IL1RN, S100A12 and diagnosis as predictors in the final model. CD276 [hazard ratio 2.42 (1.37–4.26), p=0.002] and S100A12 [hazard ratio 1.29 (1.04–1.61), p=0.022] levels were significantly associated with time to death (Supplementary Figure 6). To illustrate this effect, we plotted separately the patients with high and low values of CD276 and S100A12, as separated by the marker-specific median (Figure 5).

**Figure 5.**
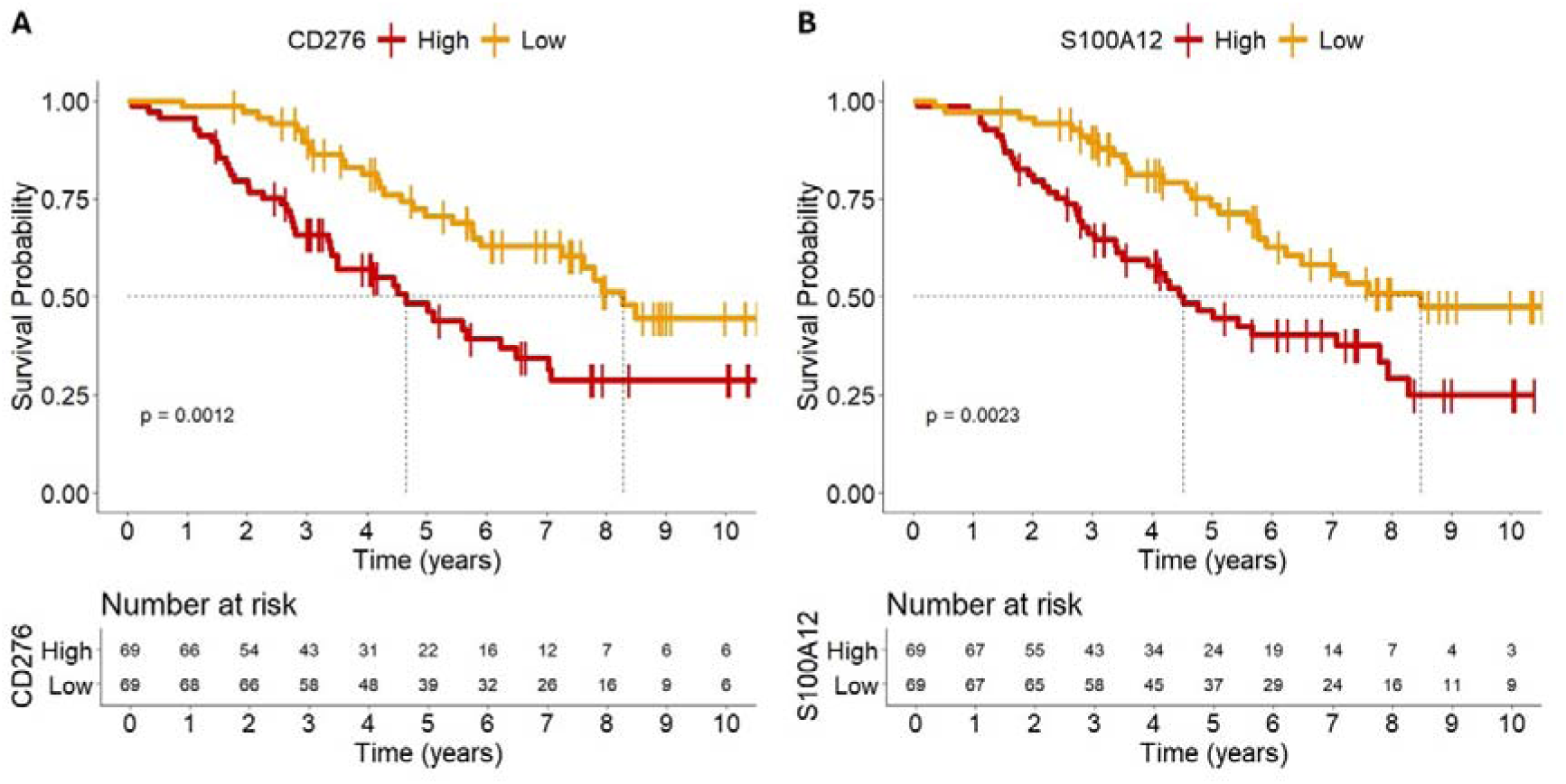
Inflammation prognostic markers. Kaplan–Meier Survival Curves of CD276 (A) and S100A12 (B) divided on the respective medians.

## 4. Discussion

Multiplex NULISA biomarkers demonstrated high sensitivity for detecting changes in serum protein levels across clinically and pathologically heterogeneous neurodegenerative disorders. There was high reliability and differential diagnostic sensitivity from the NULISA CNS panel, expanding previous evidence by (i) the inclusion of patients with sporadic FTD and PSP, and (ii) the prediction of survival. The NULISA Inflammation 250 panel detected changes in inflammatory patterns across multiple neurodegenerative diseases, with anti-inflammatory and pro-inflammatory changes, highlighting the complexity in interpreting cross-sectional plasma biomarkers in dynamic diseases.

Recent studies have shown the potential utility of the NULISA CNS panel to quantify dementia-relevant markers, including phosphorylated tau epitopes, NfL and GFAP, in blood of patients with MCI and/or AD [12–14], genetic FTD and LBD [13]. In our cohort, increased levels of serum NfL were identified across all patient groups as compared to controls, and were most elevated in the FTD and PSP cohorts. On the other hand, phosphorylated tau epitopes were the most significant markers in separating patients with AD and LBD from controls. These findings align with previous evidence with single-marker assays in similar cohorts [5–10], and suggest common AD co-pathology in the LBD cohort. Across all groups, we compared the NULISA CNS serum results with Simoa plasma assays for p-tau217, p-tau231, NfL and GFAP, finding strong correlations. This suggests the multiplex panel data can be as informative as more targeted assays, showing good consistency with the advantage of quantifying additional markers, using either on plasma or serum samples. This aligns with and expands similar evidence in cohorts of healthy volunteers and patients with AD [12,14]. Importantly, the quantification of the CNS serum proteins in our cohort was not affected by the time they have been stored in freezers at -70/- 80 □.

We also demonstrate that dementia-relevant markers from the CNS panel are prognostic, against survival. Serum NfL levels were associated with subsequent faster decline and shorter time from blood sampling to death, over and above p-tau217, p-tau231 and GFAP levels, and over and above age and diagnosis. People with FTD and PSP show the strongest associations between NfL and survival. Serum and plasma NfL levels are promising markers for disease progression and may be useful trial endpoints in patients with FTD and PSP [6,27–32]. Here, survival rate was used to test for the prognostic utility of these markers as the high clinical heterogeneity across diagnostic groups makes it difficult to capture disease severity at the time of blood collection.

Beyond the common dementia-relevant markers previously described, the NULISA CNS panel in our cohort identified lower levels of CRH in all patient groups compared to controls. CRH is an important stress hormone that may be expected to be higher in diverse disease states. Impairment of the hypothalamic pituitary adrenal (HPA) axis has been long recognised in neurodegenerative disease. At post-mortem, diminished CRH is seen across brain regions in AD [33], while CSF cortisol is elevated in patients with AD versus controls [34]. One hypothesis for reduced levels of CRH in patients is endocrine insufficiency owing to hypothalamus neurodegeneration. Alternatively, high plasma cortisol is known to occur in AD, relating to worse hippocampal atrophy and clinical progression ^35,36^; the resultant negative feedback loop may sufficiently explain the pattern of CRH being consistently higher in controls than patients. CRH is the only HPA axis protein examined in the NULISA CNS panel, thus further investigations are required to clarify the biological underpinnings of this finding. Lower CRH levels were also associated with worse survival (Figure 4), with the strongest associations being in the LBD cohort (Supplementary Figure 4). In line with our findings, CRH has emerged as a biomarker of interest in LBD, but evidence to date has been CSF-based [35,36].

Across the CNS and Inflammation panels, patients with FTD and PSP showed upregulation of many inflammation markers, as compared to controls and patients with AD. Specifically, we identified pro-inflammatory proteins oncostatin M (OSM), matrix metalloproteinase 9 (MMP9) and the potent chemo-attractant CCL7 to be elevated in PSP and FTD versus controls. We found significantly increased levels of the anti-inflammatory proteins CD276 and AREG in PSP, FTD and LBD versus controls. The anti-inflammatory IL1 receptor antagonist (IL1RN) and the neuroprotective hepatocyte growth factor (HGF) were elevated in FTD and PSP versus controls only. Although previous fluid markers confirmed abnormally high levels of inflammation in patients with FTD and PSP (see [37] for review), they have been largely confined to CSF, small case reports, limited panels and/or single markers. Our recent study, using Meso-Scale Discovery assays on serum samples, identified a pattern of pro-inflammatory cytokines upregulated in 214 patients with FTD, PSP and related conditions [30]. This inflammatory pattern, including TNF-α, TNF-R1, CSF-1, IL-17A, IL-12, IP-10, IL-6 and others, differentiated patients (all syndromes) from controls and was associated with significantly lower survival over time. Importantly, proinflammatory cytokines were associated with microglial activation in frontal and brainstem regions, as quantified by TSPO PET. The NULISA Inflammation panel provides simultaneous quantification of a larger number of inflammation markers, with better detectability for specific targets than previously used technologies [11]. In our cohort, patients with PSP or FTD over-expressed proteomic markers of inflammatory/synaptic function as compared to controls and patients with AD. The degree of dysregulation across the landscape of inflammatory protein expression (as seen in Figure 2) supports that FTD and PSP correlate with more peripheral markers of inflammation than in our cohort of AD. Further studies should investigate the inflammatory patterns on the AD spectrum with larger sample sizes, that allow the separation of patients in early and late stages of the disease.

Several targets emerged as upregulated across diagnostic groups as compared to controls in our cohort. In particular, CD276, also known as B7-H3, is primarily an anti-inflammatory or immunosuppressive molecule. This immune checkpoint plays a complex role in the immune system, but its overall effect tends to inhibit immune responses, particularly in the context of cancer [38]. Although CD276 is predominantly anti-inflammatory, in non-small cell lung cancer, higher CD276 expression has been associated with increased numbers of CD8+ T cells and plasmacytoid dendritic cells [38]. In our cohort, higher levels of serum CD276 were associated with shorter survival, or time to death from blood sampling. However, the role of CD276 in non-cancerous neurological disorders has not been investigated before and further studies would be needed to elucidate its role in these conditions. When investigating the prognostic values of inflammatory markers on survival rates, higher levels of S100A12 (S100 calcium-binding protein A12) were also associated with faster decline. S100A12 exhibits potent pro-inflammatory effects through multiple mechanisms, including acting as a strong chemoattractant for monocytes and neutrophils. It binds to receptors such as RAGE (receptor for advanced glycation end products) and TLR4 (Toll-like receptor 4), triggering intracellular signalling cascades that lead to the production of pro-inflammatory cytokines like TNF-α, IL-1β, and IL-6. Along with S100A9, S100A12 has been found to be associated with the neuropathological hallmarks of AD [39]. This suggests that S100A12 may play a role in the inflammatory processes and protein complex formation observed in AD brains.

This study has several limitations. First, patient groups were included and defined according to clinical diagnosis rather than definitive pathological confirmation, thus misdiagnosis and/ or co-pathologies may occur. Second, blood-based inflammatory markers may capture comorbidities that are not directly linked to dementia, such as co-occurring inflammatory conditions or the use of specific anti-inflammatory medication. However, these are unlikely to occur in our cohort as the contributory clinical studies excluded recent systemic infections and concurrent autoimmune diseases. Of note, there was no transdiagnostic difference in C-reactive protein, a commonly used clinical marker of non-specific systemic inflammation. Third, our cohort was predominantly white/Caucasian reflecting the ethnicity distribution of the over 65-year-old population in the UK (94% ‘white’ in the 2021 national census). Further studies are needed to test the generalisation of results to other groups and to identify environmental, genetic and socioeconomic factors that might influence proteomic changes in dementia. Fourth, serum markers were measured at a single time point, whilst longitudinal studies would be needed to test-retest reliability and for disease-related dynamics over time. We have not accounted for disease stage or clinical severity at the time of blood collection, which may explain some of the heterogeneity seen across and within disease groups. Fifth, peripheral blood may not be reflective of the environment within the CNS, and patterns discovered may vary depending on blood brain barrier integrity, which varies across pathologies. Sixth, while there is mixed evidence of temporal stability of p-tau epitopes, NfL and GFAP, it remains to be seen if the remainder of the proteins highlighted by this multiplex approach are impacted by time-of-day and circadian rhythm or other factors such as posture, exercise, diet or glymphatic clearance in sleep[40–42]. Finally, it should be noted that the correlation analyses across analytical platforms were performed using two different sample types (serum for NULISA and plasma for Simoa), which may explain modest correlations for some of the markers.

In conclusion, we have shown that a multiplex approach offers consistent results compared to more targeted single molecule arrays, while simultaneously enabling a much broader scope of analysis. The NULISA CNS and Inflammation panels identify disease specific patterns of neurodegeneration and neuroinflammation. This unique disease fingerprint offers insight into disease specific pathogenesis, with potential inferences that this dysregulation may be deleterious and amenable to drug treatment. Treatment strategies that are highly efficacious at removing amyloid from the brain have shown only moderate clinical efficacy, thus it appears that alternative or indeed dual treatment strategies may be needed to tackle what are complex, multidimensional pathologies. This multiplex approach identified NfL, CRH, CD276 and S100A12 as useful prognostic markers across neurodegenerative diseases. Multiplex assay approach may offer scalable outcome measures for clinical trials in neurodegenerative diseases.

## Abbreviations

(NfL / NEFL): neurofilament light chain;
NULISA): NUcleic acid Linked Immuno-Sandwich Assay;
(NPQ): NULISA Protein Quantification;
(SIMOA): Quanterix Single molecule array

## Acknowledgments

We thank our participant volunteers for their participation in this study, thank the National Institute for Health Research (NIHR) Cambridge BioResource centre staff, and the research nurses for their contribution, and the East Anglia Dementias and Neurodegenerative Diseases Research Network (DeNDRoN) for help with subject recruitment.

For the purpose of open access, the authors have applied a Creative Commons Attribution (CC BY) license to any Author Accepted Manuscript version arising from this submission. This work is licensed under a Creative Commons Attribution 4.0 International License.

