## Supplementary Material for "Novel blood-based proteomic signatures across multiple neurodegenerative diseases"

Cambridge CB2 0SZ, UK

Supplementary Figure 1. Associations between dementia-relevant NULISA CNS markers in serum and sample storage time in freezers at -70 ºC.

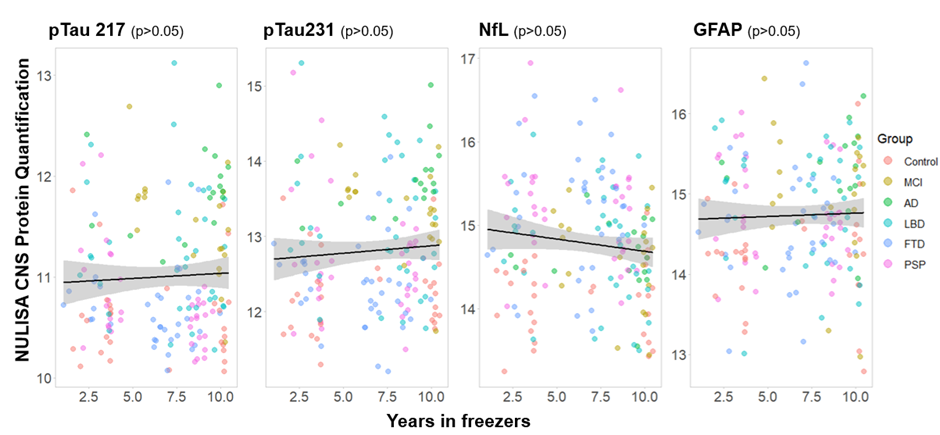

Supplementary Figure 2. Comparisons between patient groups on CNS markers. Orange dots represent comparisons with p<0.05, and red dots with p<0.05 corrected for multiple comparisons using the false discovery rate (FDR)).

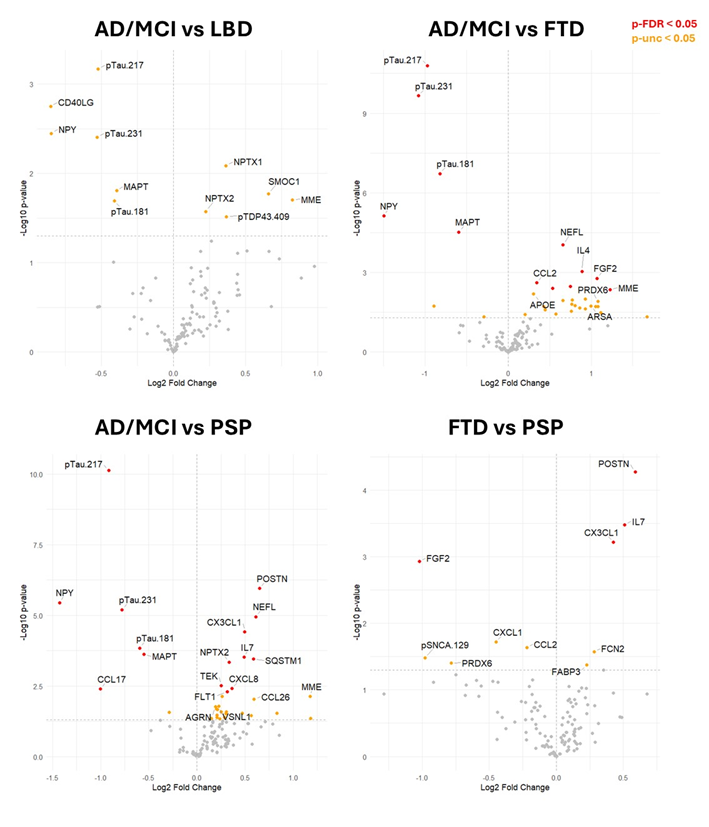

Supplementary Figure 3. Comparisons between patient groups on inflammation markers. Orange dots represent comparisons with p<0.05, and red dots with p<0.05 corrected for multiple comparisons.

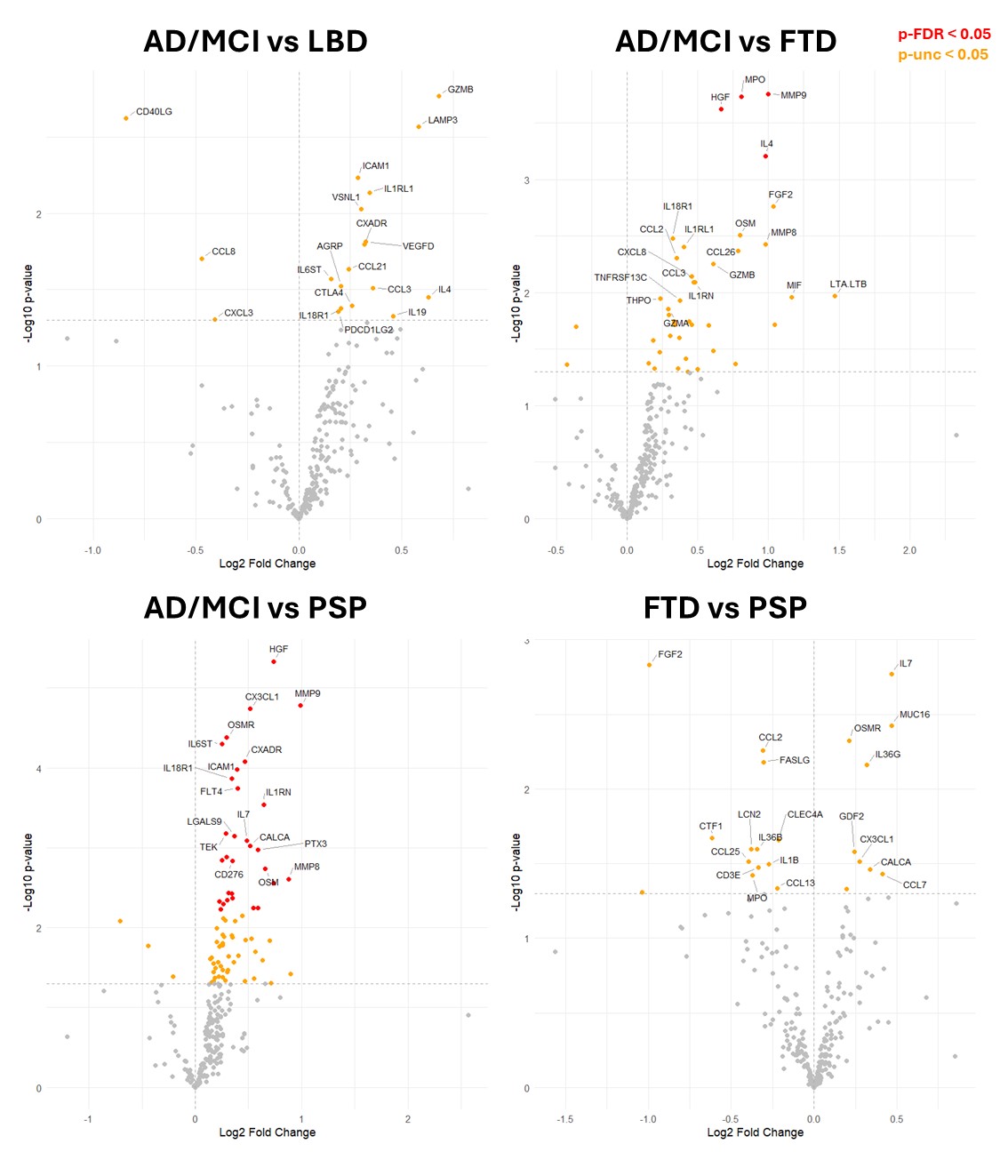

Supplementary Figure 4*.* Prognostic markers by diagnosis: A) Kaplan–Meier Survival Curve of NfL divided on the median; B) Kaplan–Meier Survival Curve of CRH divided on the median.

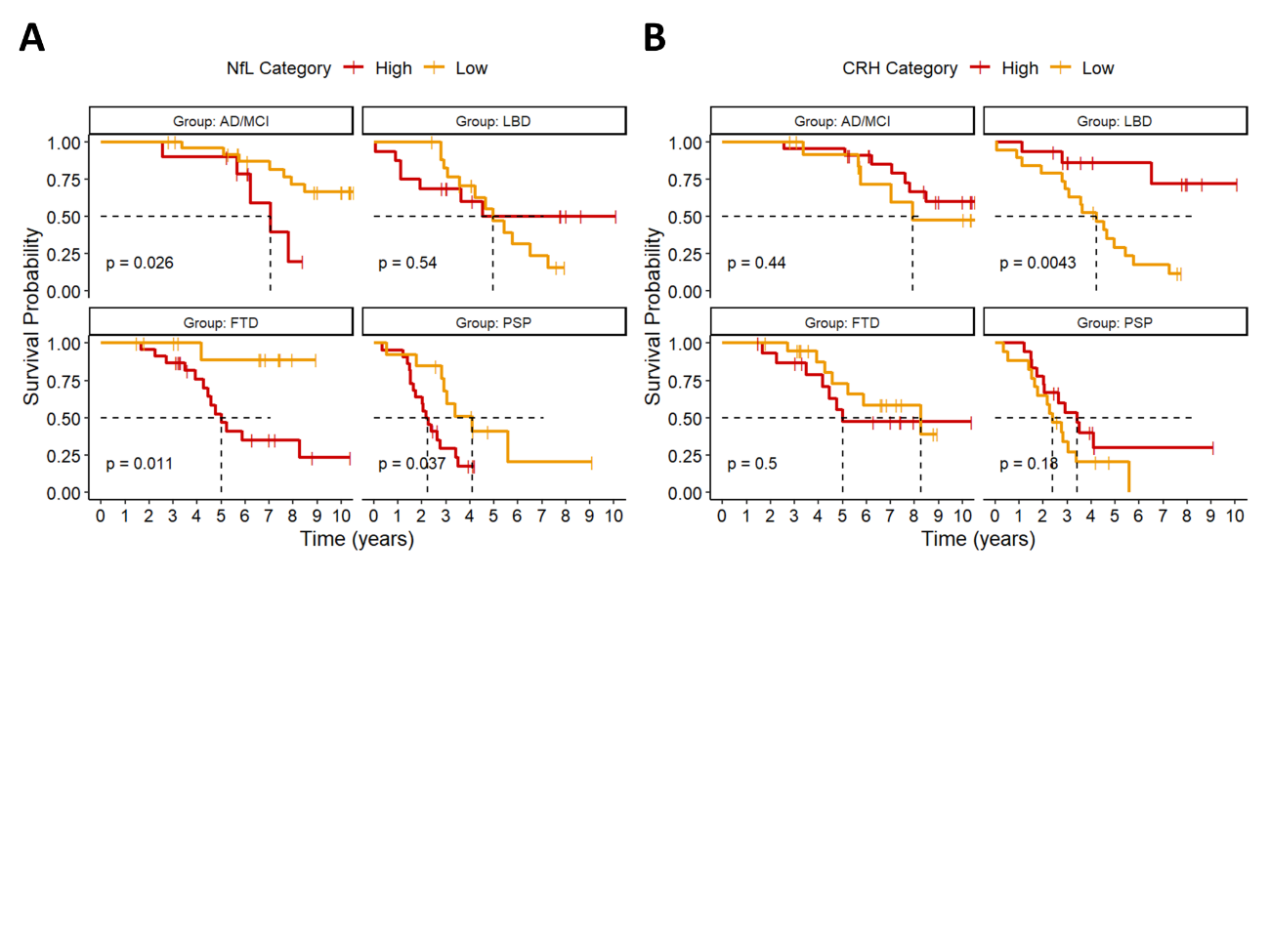

Supplementary Figure 5. Comparison across all patients vs controls on inflammation markers. Orange dots represent comparisons with p<0.05, and red dots with p<0.05 corrected for multiple comparisons. The right panel represent markers that are upregulated in patients as compared to controls, while the left panel represents markers that have lower levels in patients.

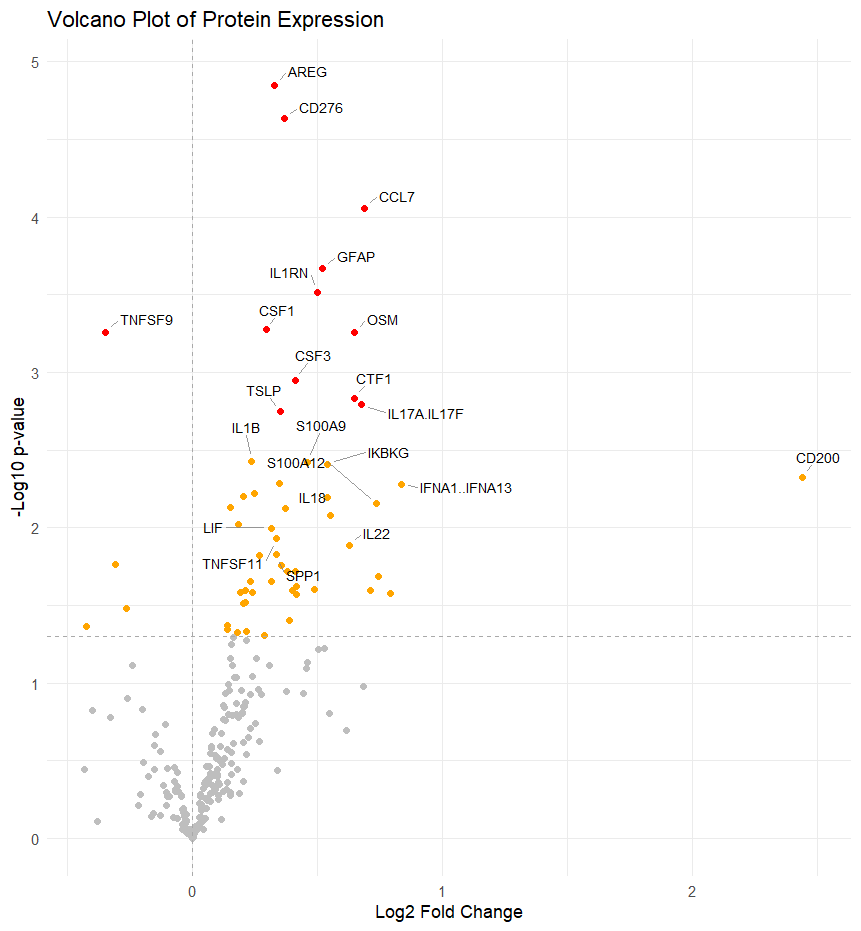

*Supplementary Figure 6. Prognostic markers on the Inflammation panel. Final model selected by the backward stepwise selection of predictors in the survival analysis including inflammatory markers, age and diagnosis. For each variable the hazard ratio on survival is plotted.*

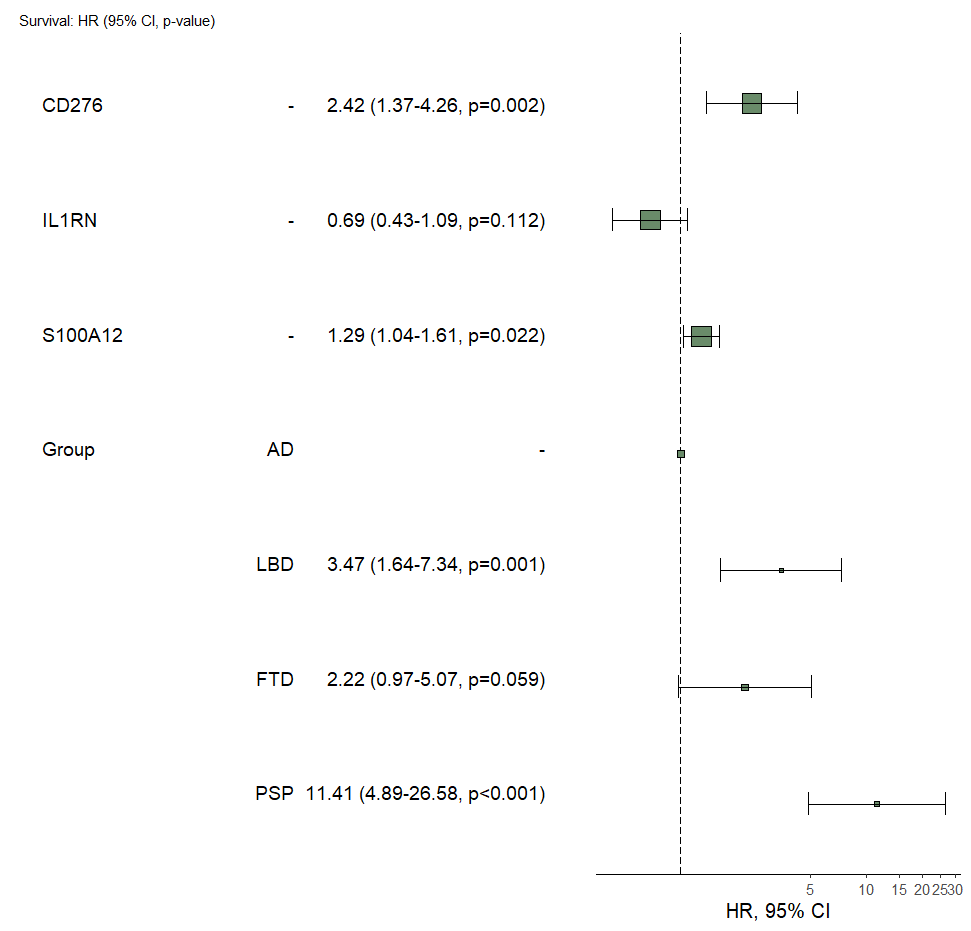

Supplementary Table 1. List of targets in NULISAseq CNS panel.

| **Target Name** | **Panel** |
| --- | --- |
| ACHE | NULISAseq CNS Panel |
| AGRN | NULISAseq CNS Panel |
| ANXA5 | NULISAseq CNS Panel |
| APOE | NULISAseq CNS Panel |
| ARSA | NULISAseq CNS Panel |
| Aβ38 | NULISAseq CNS Panel |
| Aβ40 | NULISAseq CNS Panel |
| Aβ42 | NULISAseq CNS Panel |
| BACE1 | NULISAseq CNS Panel |
| BASP1 | NULISAseq CNS Panel |
| BDNF | NULISAseq CNS Panel |
| CALB2 | NULISAseq CNS Panel |
| CCL11 | NULISAseq CNS Panel |
| CCL13 | NULISAseq CNS Panel |
| CCL17 | NULISAseq CNS Panel |
| CCL2 | NULISAseq CNS Panel |
| CCL22 | NULISAseq CNS Panel |
| CCL26 | NULISAseq CNS Panel |
| CCL3 | NULISAseq CNS Panel |
| CCL4 | NULISAseq CNS Panel |
| CD40LG | NULISAseq CNS Panel |
| CD63 | NULISAseq CNS Panel |
| CHI3L1 | NULISAseq CNS Panel |
| CHIT1 | NULISAseq CNS Panel |
| CNTN2 | NULISAseq CNS Panel |
| CRH | NULISAseq CNS Panel |
| CRP | NULISAseq CNS Panel |
| CSF2 | NULISAseq CNS Panel |
| CST3 | NULISAseq CNS Panel |
| CX3CL1 | NULISAseq CNS Panel |
| CXCL1 | NULISAseq CNS Panel |
| CXCL10 | NULISAseq CNS Panel |
| CXCL8 | NULISAseq CNS Panel |
| ENO2 | NULISAseq CNS Panel |
| FABP3 | NULISAseq CNS Panel |
| FCN2 | NULISAseq CNS Panel |
| FGF2 | NULISAseq CNS Panel |
| FLT1 | NULISAseq CNS Panel |
| FOLR1 | NULISAseq CNS Panel |
| GDF15 | NULISAseq CNS Panel |
| GDI1 | NULISAseq CNS Panel |
| GDNF | NULISAseq CNS Panel |
| GFAP | NULISAseq CNS Panel |
| GOT1 | NULISAseq CNS Panel |
| HBA1 | NULISAseq CNS Panel |
| HTT | NULISAseq CNS Panel |
| ICAM1 | NULISAseq CNS Panel |
| IFNG | NULISAseq CNS Panel |
| IGF1R | NULISAseq CNS Panel |
| IGFBP7 | NULISAseq CNS Panel |
| IL10 | NULISAseq CNS Panel |
| IL12p70 | NULISAseq CNS Panel |
| IL13 | NULISAseq CNS Panel |
| IL15 | NULISAseq CNS Panel |
| IL16 | NULISAseq CNS Panel |
| IL17A | NULISAseq CNS Panel |
| IL18 | NULISAseq CNS Panel |
| IL1B | NULISAseq CNS Panel |
| IL2 | NULISAseq CNS Panel |
| IL33 | NULISAseq CNS Panel |
| IL4 | NULISAseq CNS Panel |
| IL5 | NULISAseq CNS Panel |
| IL6 | NULISAseq CNS Panel |
| IL6R | NULISAseq CNS Panel |
| IL7 | NULISAseq CNS Panel |
| IL9 | NULISAseq CNS Panel |
| KDR | NULISAseq CNS Panel |
| KLK6 | NULISAseq CNS Panel |
| MAPT | NULISAseq CNS Panel |
| MDH1 | NULISAseq CNS Panel |
| MME | NULISAseq CNS Panel |
| MSLN | NULISAseq CNS Panel |
| NEFH | NULISAseq CNS Panel |
| NEFL (or NfL) | NULISAseq CNS Panel |
| NGF | NULISAseq CNS Panel |
| NPTX1 | NULISAseq CNS Panel |
| NPTX2 | NULISAseq CNS Panel |
| NPTXR | NULISAseq CNS Panel |
| NPY | NULISAseq CNS Panel |
| NRGN | NULISAseq CNS Panel |
| Oligo-SNCA | NULISAseq CNS Panel |
| PARK7 | NULISAseq CNS Panel |
| PDGFRB | NULISAseq CNS Panel |
| PDLIM5 | NULISAseq CNS Panel |
| PGF | NULISAseq CNS Panel |
| PGK1 | NULISAseq CNS Panel |
| POSTN | NULISAseq CNS Panel |
| PRDX6 | NULISAseq CNS Panel |
| PSEN1 | NULISAseq CNS Panel |
| pSNCA-129 | NULISAseq CNS Panel |
| p-tau-181 | NULISAseq CNS Panel |
| p-tau-217 | NULISAseq CNS Panel |
| p-tau-231 | NULISAseq CNS Panel |
| pTDP43-409 | NULISAseq CNS Panel |
| PTN | NULISAseq CNS Panel |
| REST | NULISAseq CNS Panel |
| RUVBL2 | NULISAseq CNS Panel |
| S100A12 | NULISAseq CNS Panel |
| S100B | NULISAseq CNS Panel |
| SAA1 | NULISAseq CNS Panel |
| SFRP1 | NULISAseq CNS Panel |
| SFTPD | NULISAseq CNS Panel |
| SLIT2 | NULISAseq CNS Panel |
| SMOC1 | NULISAseq CNS Panel |
| SNAP25 | NULISAseq CNS Panel |
| SNCA | NULISAseq CNS Panel |
| SNCB | NULISAseq CNS Panel |
| SOD1 | NULISAseq CNS Panel |
| SQSTM1 | NULISAseq CNS Panel |
| TAFA5 | NULISAseq CNS Panel |
| TARDBP | NULISAseq CNS Panel |
| TEK | NULISAseq CNS Panel |
| TIMP3 | NULISAseq CNS Panel |
| TNF | NULISAseq CNS Panel |
| TREM1 | NULISAseq CNS Panel |
| TREM2 | NULISAseq CNS Panel |
| UCHL1 | NULISAseq CNS Panel |
| VCAM1 | NULISAseq CNS Panel |
| VEGFA | NULISAseq CNS Panel |
| VEGFD | NULISAseq CNS Panel |
| VGF | NULISAseq CNS Panel |
| VSNL1 | NULISAseq CNS Panel |
| YWHAZ | NULISAseq CNS Panel |

Supplementary Table 2. List of targets in NULISAseq Inflammation panel.

| **Target Name** | **Panel** |
| --- | --- |
| AGER | NULISAseq Inflammation Panel |
| AGRP | NULISAseq Inflammation Panel |
| ANGPT1 | NULISAseq Inflammation Panel |
| ANGPT2 | NULISAseq Inflammation Panel |
| ANXA1 | NULISAseq Inflammation Panel |
| AREG | NULISAseq Inflammation Panel |
| BDNF | NULISAseq Inflammation Panel |
| BMP7 | NULISAseq Inflammation Panel |
| BST2 | NULISAseq Inflammation Panel |
| C1QA | NULISAseq Inflammation Panel |
| CALCA | NULISAseq Inflammation Panel |
| CCL1 | NULISAseq Inflammation Panel |
| CCL11 | NULISAseq Inflammation Panel |
| CCL13 | NULISAseq Inflammation Panel |
| CCL14 | NULISAseq Inflammation Panel |
| CCL15 | NULISAseq Inflammation Panel |
| CCL16 | NULISAseq Inflammation Panel |
| CCL17 | NULISAseq Inflammation Panel |
| CCL19 | NULISAseq Inflammation Panel |
| CCL2 | NULISAseq Inflammation Panel |
| CCL20 | NULISAseq Inflammation Panel |
| CCL21 | NULISAseq Inflammation Panel |
| CCL22 | NULISAseq Inflammation Panel |
| CCL23 | NULISAseq Inflammation Panel |
| CCL24 | NULISAseq Inflammation Panel |
| CCL25 | NULISAseq Inflammation Panel |
| CCL26 | NULISAseq Inflammation Panel |
| CCL27 | NULISAseq Inflammation Panel |
| CCL28 | NULISAseq Inflammation Panel |
| CCL3 | NULISAseq Inflammation Panel |
| CCL4 | NULISAseq Inflammation Panel |
| CCL5 | NULISAseq Inflammation Panel |
| CCL7 | NULISAseq Inflammation Panel |
| CCL8 | NULISAseq Inflammation Panel |
| CD200 | NULISAseq Inflammation Panel |
| CD200R1 | NULISAseq Inflammation Panel |
| CD27 | NULISAseq Inflammation Panel |
| CD274 | NULISAseq Inflammation Panel |
| CD276 | NULISAseq Inflammation Panel |
| CD3E | NULISAseq Inflammation Panel |
| CD4 | NULISAseq Inflammation Panel |
| CD40 | NULISAseq Inflammation Panel |
| CD40LG | NULISAseq Inflammation Panel |
| CD46 | NULISAseq Inflammation Panel |
| CD70 | NULISAseq Inflammation Panel |
| CD80 | NULISAseq Inflammation Panel |
| CD83 | NULISAseq Inflammation Panel |
| CD93 | NULISAseq Inflammation Panel |
| CEACAM5 | NULISAseq Inflammation Panel |
| CHI3L1 | NULISAseq Inflammation Panel |
| CLEC4A | NULISAseq Inflammation Panel |
| CNTF | NULISAseq Inflammation Panel |
| CRP | NULISAseq Inflammation Panel |
| CSF1 | NULISAseq Inflammation Panel |
| CSF1R | NULISAseq Inflammation Panel |
| CSF2 | NULISAseq Inflammation Panel |
| CSF2RB | NULISAseq Inflammation Panel |
| CSF3 | NULISAseq Inflammation Panel |
| CSF3R | NULISAseq Inflammation Panel |
| CST7 | NULISAseq Inflammation Panel |
| CTF1 | NULISAseq Inflammation Panel |
| CTLA4 | NULISAseq Inflammation Panel |
| CTSS | NULISAseq Inflammation Panel |
| CX3CL1 | NULISAseq Inflammation Panel |
| CXADR | NULISAseq Inflammation Panel |
| CXCL1 | NULISAseq Inflammation Panel |
| CXCL10 | NULISAseq Inflammation Panel |
| CXCL11 | NULISAseq Inflammation Panel |
| CXCL12 | NULISAseq Inflammation Panel |
| CXCL13 | NULISAseq Inflammation Panel |
| CXCL14 | NULISAseq Inflammation Panel |
| CXCL16 | NULISAseq Inflammation Panel |
| CXCL2 | NULISAseq Inflammation Panel |
| CXCL3 | NULISAseq Inflammation Panel |
| CXCL5 | NULISAseq Inflammation Panel |
| CXCL6 | NULISAseq Inflammation Panel |
| CXCL8 | NULISAseq Inflammation Panel |
| CXCL9 | NULISAseq Inflammation Panel |
| EGF | NULISAseq Inflammation Panel |
| EPO | NULISAseq Inflammation Panel |
| FASLG | NULISAseq Inflammation Panel |
| FGF19 | NULISAseq Inflammation Panel |
| FGF2 | NULISAseq Inflammation Panel |
| FGF21 | NULISAseq Inflammation Panel |
| FGF23 | NULISAseq Inflammation Panel |
| FLT1 | NULISAseq Inflammation Panel |
| FLT3LG | NULISAseq Inflammation Panel |
| FLT4 | NULISAseq Inflammation Panel |
| FTH1 | NULISAseq Inflammation Panel |
| FURIN | NULISAseq Inflammation Panel |
| GDF15 | NULISAseq Inflammation Panel |
| GDF2 | NULISAseq Inflammation Panel |
| GFAP | NULISAseq Inflammation Panel |
| GRN | NULISAseq Inflammation Panel |
| GZMA | NULISAseq Inflammation Panel |
| GZMB | NULISAseq Inflammation Panel |
| HAVCR1 | NULISAseq Inflammation Panel |
| HGF | NULISAseq Inflammation Panel |
| HLA-DRA | NULISAseq Inflammation Panel |
| ICAM1 | NULISAseq Inflammation Panel |
| ICOSLG | NULISAseq Inflammation Panel |
| IFNA1; IFNA13 | NULISAseq Inflammation Panel |
| IFNA2 | NULISAseq Inflammation Panel |
| IFNB1 | NULISAseq Inflammation Panel |
| IFNG | NULISAseq Inflammation Panel |
| IFNL1 | NULISAseq Inflammation Panel |
| IFNL2; IFNL3 | NULISAseq Inflammation Panel |
| IFNW1 | NULISAseq Inflammation Panel |
| IKBKG | NULISAseq Inflammation Panel |
| IL10 | NULISAseq Inflammation Panel |
| IL10RB | NULISAseq Inflammation Panel |
| IL11 | NULISAseq Inflammation Panel |
| IL12B | NULISAseq Inflammation Panel |
| IL12p70 | NULISAseq Inflammation Panel |
| IL12RB1 | NULISAseq Inflammation Panel |
| IL13 | NULISAseq Inflammation Panel |
| IL13RA2 | NULISAseq Inflammation Panel |
| IL15 | NULISAseq Inflammation Panel |
| IL15RA | NULISAseq Inflammation Panel |
| IL16 | NULISAseq Inflammation Panel |
| IL17A | NULISAseq Inflammation Panel |
| IL17A\|IL17F | NULISAseq Inflammation Panel |
| IL17B | NULISAseq Inflammation Panel |
| IL17C | NULISAseq Inflammation Panel |
| IL17F | NULISAseq Inflammation Panel |
| IL17RA | NULISAseq Inflammation Panel |
| IL17RB | NULISAseq Inflammation Panel |
| IL18 | NULISAseq Inflammation Panel |
| IL18BP | NULISAseq Inflammation Panel |
| IL18R1 | NULISAseq Inflammation Panel |
| IL19 | NULISAseq Inflammation Panel |
| IL1B | NULISAseq Inflammation Panel |
| IL1R1 | NULISAseq Inflammation Panel |
| IL1R2 | NULISAseq Inflammation Panel |
| IL1RL1 | NULISAseq Inflammation Panel |
| IL1RN | NULISAseq Inflammation Panel |
| IL2 | NULISAseq Inflammation Panel |
| IL20 | NULISAseq Inflammation Panel |
| IL22 | NULISAseq Inflammation Panel |
| IL23 | NULISAseq Inflammation Panel |
| IL24 | NULISAseq Inflammation Panel |
| IL27 | NULISAseq Inflammation Panel |
| IL2RA | NULISAseq Inflammation Panel |
| IL2RB | NULISAseq Inflammation Panel |
| IL32 | NULISAseq Inflammation Panel |
| IL33 | NULISAseq Inflammation Panel |
| IL34 | NULISAseq Inflammation Panel |
| IL36A | NULISAseq Inflammation Panel |
| IL36B | NULISAseq Inflammation Panel |
| IL36G | NULISAseq Inflammation Panel |
| IL3RA | NULISAseq Inflammation Panel |
| IL4 | NULISAseq Inflammation Panel |
| IL4R | NULISAseq Inflammation Panel |
| IL5 | NULISAseq Inflammation Panel |
| IL5RA | NULISAseq Inflammation Panel |
| IL6 | NULISAseq Inflammation Panel |
| IL6R | NULISAseq Inflammation Panel |
| IL6ST | NULISAseq Inflammation Panel |
| IL7 | NULISAseq Inflammation Panel |
| IL7R | NULISAseq Inflammation Panel |
| IL9 | NULISAseq Inflammation Panel |
| IRAK4 | NULISAseq Inflammation Panel |
| KDR | NULISAseq Inflammation Panel |
| KITLG | NULISAseq Inflammation Panel |
| KLRK1 | NULISAseq Inflammation Panel |
| KNG1 | NULISAseq Inflammation Panel |
| LAG3 | NULISAseq Inflammation Panel |
| LAMP3 | NULISAseq Inflammation Panel |
| LCN2 | NULISAseq Inflammation Panel |
| LGALS9 | NULISAseq Inflammation Panel |
| LIF | NULISAseq Inflammation Panel |
| LILRB2 | NULISAseq Inflammation Panel |
| LTA | NULISAseq Inflammation Panel |
| LTA\|LTB | NULISAseq Inflammation Panel |
| MERTK | NULISAseq Inflammation Panel |
| MICA | NULISAseq Inflammation Panel |
| MICB | NULISAseq Inflammation Panel |
| MIF | NULISAseq Inflammation Panel |
| MMP1 | NULISAseq Inflammation Panel |
| MMP12 | NULISAseq Inflammation Panel |
| MMP3 | NULISAseq Inflammation Panel |
| MMP8 | NULISAseq Inflammation Panel |
| MMP9 | NULISAseq Inflammation Panel |
| MPO | NULISAseq Inflammation Panel |
| MUC16 | NULISAseq Inflammation Panel |
| NAMPT | NULISAseq Inflammation Panel |
| NCR1 | NULISAseq Inflammation Panel |
| NGF | NULISAseq Inflammation Panel |
| NTF3 | NULISAseq Inflammation Panel |
| OSM | NULISAseq Inflammation Panel |
| OSMR | NULISAseq Inflammation Panel |
| PDCD1 | NULISAseq Inflammation Panel |
| PDCD1LG2 | NULISAseq Inflammation Panel |
| PDGFA | NULISAseq Inflammation Panel |
| PDGFB | NULISAseq Inflammation Panel |
| PGF | NULISAseq Inflammation Panel |
| PTX3 | NULISAseq Inflammation Panel |
| S100A12 | NULISAseq Inflammation Panel |
| S100A9 | NULISAseq Inflammation Panel |
| SCG2 | NULISAseq Inflammation Panel |
| SDC1 | NULISAseq Inflammation Panel |
| SELE | NULISAseq Inflammation Panel |
| SELP | NULISAseq Inflammation Panel |
| SIRPA | NULISAseq Inflammation Panel |
| SLAMF1 | NULISAseq Inflammation Panel |
| SPP1 | NULISAseq Inflammation Panel |
| TAFA5 | NULISAseq Inflammation Panel |
| TEK | NULISAseq Inflammation Panel |
| TGFB1 | NULISAseq Inflammation Panel |
| TGFB3 | NULISAseq Inflammation Panel |
| THBS2 | NULISAseq Inflammation Panel |
| THPO | NULISAseq Inflammation Panel |
| TIMP1 | NULISAseq Inflammation Panel |
| TIMP2 | NULISAseq Inflammation Panel |
| TLR3 | NULISAseq Inflammation Panel |
| TNF | NULISAseq Inflammation Panel |
| TNFRSF11A | NULISAseq Inflammation Panel |
| TNFRSF11B | NULISAseq Inflammation Panel |
| TNFRSF13B | NULISAseq Inflammation Panel |
| TNFRSF13C | NULISAseq Inflammation Panel |
| TNFRSF14 | NULISAseq Inflammation Panel |
| TNFRSF17 | NULISAseq Inflammation Panel |
| TNFRSF18 | NULISAseq Inflammation Panel |
| TNFRSF1A | NULISAseq Inflammation Panel |
| TNFRSF1B | NULISAseq Inflammation Panel |
| TNFRSF21 | NULISAseq Inflammation Panel |
| TNFRSF4 | NULISAseq Inflammation Panel |
| TNFRSF8 | NULISAseq Inflammation Panel |
| TNFRSF9 | NULISAseq Inflammation Panel |
| TNFSF10 | NULISAseq Inflammation Panel |
| TNFSF11 | NULISAseq Inflammation Panel |
| TNFSF12 | NULISAseq Inflammation Panel |
| TNFSF13 | NULISAseq Inflammation Panel |
| TNFSF14 | NULISAseq Inflammation Panel |
| TNFSF15 | NULISAseq Inflammation Panel |
| TNFSF18 | NULISAseq Inflammation Panel |
| TNFSF4 | NULISAseq Inflammation Panel |
| TNFSF8 | NULISAseq Inflammation Panel |
| TNFSF9 | NULISAseq Inflammation Panel |
| TREM1 | NULISAseq Inflammation Panel |
| TREM2 | NULISAseq Inflammation Panel |
| TSLP | NULISAseq Inflammation Panel |
| VCAM1 | NULISAseq Inflammation Panel |
| VEGFA | NULISAseq Inflammation Panel |
| VEGFC | NULISAseq Inflammation Panel |
| VEGFD | NULISAseq Inflammation Panel |
| VSNL1 | NULISAseq Inflammation Panel |
| VSTM1 | NULISAseq Inflammation Panel |
| WNT16 | NULISAseq Inflammation Panel |
| WNT7A | NULISAseq Inflammation Panel |

Supplementary Table 3. LIMMA full results on NULISA CNS panel markers, comparing each diagnostic group with controls. Uncorrected p values < 0.05 are highlighted in yellow, while adjusted p values < 0.05 are highlighted in red.

|  | **Controls vs AD/MCI** | | | | **Controls vs LBD** | | | | **Controls vs FTD** | | | | **Controls vs PSP** | | | |
| --- | --- | --- | --- | --- | --- | --- | --- | --- | --- | --- | --- | --- | --- | --- | --- | --- |
| Marker | **logFC** | **t** | **p** | **adj p** | **logFC** | **t** | **p** | **adj p** | **logFC** | **t** | **p** | **adj p** | **logFC** | **t** | **p** | **adj p** |
| **A-38** | -0.21 | -0.86 | 0.39 | 0.81 | -0.72 | -2.45 | 0.02 | 0.25 | -0.42 | -1.46 | 0.15 | 0.38 | -0.38 | -1.81 | 0.08 | 0.34 |
| **A-40** | -0.17 | -0.88 | 0.38 | 0.81 | -0.49 | -1.99 | 0.05 | 0.29 | -0.23 | -0.92 | 0.36 | 0.65 | -0.24 | -1.63 | 0.11 | 0.41 |
| **A-42** | -0.31 | -2.03 | 0.05 | 0.32 | -0.19 | -1.07 | 0.29 | 0.64 | -0.47 | -3.04 | 0.00 | 0.04 | -0.09 | -0.59 | 0.56 | 0.76 |
| **ACHE** | 0.28 | 2.24 | 0.03 | 0.28 | 0.13 | 0.80 | 0.42 | 0.74 | 0.06 | 0.49 | 0.62 | 0.85 | -0.08 | -0.79 | 0.43 | 0.70 |
| **AGRN** | -0.10 | -1.13 | 0.26 | 0.69 | -0.03 | -0.26 | 0.80 | 0.94 | -0.02 | -0.29 | 0.77 | 0.96 | 0.07 | 0.92 | 0.36 | 0.65 |
| **ANXA5** | 0.18 | 0.82 | 0.41 | 0.82 | 0.32 | 1.08 | 0.29 | 0.64 | 0.29 | 1.31 | 0.20 | 0.45 | 0.52 | 1.52 | 0.13 | 0.46 |
| **APOE** | -0.03 | -0.31 | 0.76 | 0.96 | 0.04 | 0.30 | 0.76 | 0.94 | 0.28 | 2.21 | 0.03 | 0.14 | 0.08 | 0.71 | 0.48 | 0.74 |
| **ARSA** | 0.00 | -0.01 | 0.99 | 0.99 | 0.66 | 1.98 | 0.05 | 0.29 | 1.01 | 2.94 | 0.00 | 0.04 | 0.31 | 1.10 | 0.28 | 0.58 |
| **BACE1** | 0.00 | 0.05 | 0.96 | 0.97 | 0.02 | 0.25 | 0.80 | 0.94 | -0.06 | -0.58 | 0.56 | 0.85 | 0.02 | 0.27 | 0.79 | 0.84 |
| **BASP1** | -0.21 | -0.42 | 0.68 | 0.96 | 0.08 | 0.18 | 0.86 | 0.96 | -0.81 | -2.39 | 0.02 | 0.11 | -0.48 | -1.18 | 0.24 | 0.54 |
| **BDNF** | 0.01 | 0.14 | 0.89 | 0.97 | -0.17 | -1.62 | 0.11 | 0.44 | -0.04 | -0.52 | 0.60 | 0.85 | 0.02 | 0.32 | 0.75 | 0.84 |
| **CALB2** | 0.06 | 0.63 | 0.53 | 0.88 | 0.18 | 1.26 | 0.21 | 0.62 | 0.05 | 0.53 | 0.60 | 0.85 | 0.25 | 2.54 | 0.01 | 0.11 |
| **CCL11** | 0.22 | 1.80 | 0.08 | 0.38 | 0.15 | 1.13 | 0.26 | 0.64 | 0.15 | 1.26 | 0.21 | 0.46 | 0.14 | 1.31 | 0.20 | 0.51 |
| **CCL13** | 0.32 | 2.13 | 0.04 | 0.29 | 0.20 | 1.33 | 0.19 | 0.58 | 0.25 | 2.03 | 0.05 | 0.17 | 0.09 | 0.65 | 0.52 | 0.75 |
| **CCL17** | 0.74 | 2.24 | 0.03 | 0.28 | 0.13 | 0.33 | 0.74 | 0.94 | -0.05 | -0.15 | 0.88 | 0.98 | -0.28 | -0.79 | 0.43 | 0.70 |
| **CCL2** | -0.03 | -0.29 | 0.77 | 0.96 | 0.05 | 0.35 | 0.73 | 0.93 | 0.30 | 2.98 | 0.00 | 0.04 | 0.09 | 0.95 | 0.35 | 0.64 |
| **CCL22** | -0.43 | -2.23 | 0.03 | 0.28 | -0.46 | -2.28 | 0.03 | 0.25 | -0.30 | -1.65 | 0.10 | 0.30 | -0.19 | -1.04 | 0.30 | 0.61 |
| **CCL26** | -0.69 | -2.14 | 0.04 | 0.29 | -0.36 | -1.18 | 0.24 | 0.64 | 0.07 | 0.27 | 0.78 | 0.96 | -0.13 | -0.54 | 0.59 | 0.77 |
| **CCL3** | -0.39 | -1.80 | 0.08 | 0.38 | -0.04 | -0.15 | 0.88 | 0.96 | 0.19 | 0.81 | 0.42 | 0.72 | -0.07 | -0.33 | 0.74 | 0.84 |
| **CCL4** | -0.27 | -1.10 | 0.28 | 0.71 | -0.08 | -0.29 | 0.77 | 0.94 | -0.19 | -0.73 | 0.47 | 0.77 | -0.14 | -0.61 | 0.55 | 0.75 |
| **CD40LG** | 0.44 | 2.15 | 0.04 | 0.29 | -0.54 | -1.85 | 0.07 | 0.32 | 0.32 | 1.46 | 0.15 | 0.38 | 0.03 | 0.14 | 0.89 | 0.92 |
| **CD63** | 0.10 | 0.95 | 0.35 | 0.77 | 0.10 | 0.85 | 0.40 | 0.74 | 0.18 | 1.88 | 0.07 | 0.22 | 0.06 | 0.61 | 0.54 | 0.75 |
| **CHI3L1** | -0.03 | -0.13 | 0.89 | 0.97 | 0.51 | 1.92 | 0.06 | 0.30 | 0.39 | 1.93 | 0.06 | 0.21 | 0.42 | 2.22 | 0.03 | 0.19 |
| **CHIT1** | -1.12 | -1.78 | 0.08 | 0.38 | -0.12 | -0.53 | 0.60 | 0.89 | -0.13 | -0.42 | 0.68 | 0.90 | 0.05 | 0.27 | 0.79 | 0.84 |
| **CNTN2** | -0.28 | -1.25 | 0.22 | 0.65 | 0.07 | 0.24 | 0.81 | 0.94 | -0.08 | -0.36 | 0.72 | 0.92 | 0.12 | 0.55 | 0.58 | 0.76 |
| **CRH** | -0.64 | -2.57 | 0.01 | 0.19 | -0.95 | -3.08 | 0.00 | 0.10 | -0.89 | -3.49 | 0.00 | 0.02 | -0.77 | -2.86 | 0.01 | 0.08 |
| **CRP** | -0.22 | -0.53 | 0.60 | 0.89 | 0.18 | 0.46 | 0.64 | 0.90 | 0.20 | 0.50 | 0.62 | 0.85 | 0.43 | 1.16 | 0.25 | 0.54 |
| **CSF2** | -0.19 | -1.51 | 0.14 | 0.53 | -0.06 | -0.35 | 0.73 | 0.93 | -0.04 | -0.33 | 0.74 | 0.93 | -0.02 | -0.12 | 0.90 | 0.92 |
| **CST3** | -0.02 | -0.26 | 0.79 | 0.96 | 0.10 | 1.17 | 0.25 | 0.64 | 0.03 | 0.50 | 0.62 | 0.85 | 0.09 | 1.64 | 0.11 | 0.41 |
| **CX3CL1** | -0.12 | -1.07 | 0.29 | 0.72 | -0.06 | -0.50 | 0.62 | 0.90 | -0.04 | -0.34 | 0.74 | 0.93 | 0.38 | 2.89 | 0.01 | 0.08 |
| **CXCL1** | -0.08 | -0.45 | 0.66 | 0.95 | -0.20 | -1.08 | 0.28 | 0.64 | 0.43 | 1.89 | 0.06 | 0.22 | -0.04 | -0.28 | 0.78 | 0.84 |
| **CXCL10** | 0.33 | 1.78 | 0.08 | 0.38 | 0.51 | 2.55 | 0.01 | 0.24 | 0.49 | 2.57 | 0.01 | 0.08 | 0.48 | 2.81 | 0.01 | 0.08 |
| **CXCL8** | -0.07 | -0.53 | 0.60 | 0.89 | -0.07 | -0.48 | 0.63 | 0.90 | 0.53 | 2.76 | 0.01 | 0.05 | 0.28 | 2.07 | 0.04 | 0.25 |
| **ENO2** | 0.20 | 1.22 | 0.23 | 0.65 | 0.18 | 1.15 | 0.25 | 0.64 | 0.26 | 1.81 | 0.07 | 0.24 | 0.20 | 1.43 | 0.16 | 0.50 |
| **FABP3** | 0.06 | 0.50 | 0.62 | 0.91 | 0.16 | 1.07 | 0.29 | 0.64 | 0.01 | 0.09 | 0.93 | 0.99 | 0.30 | 2.73 | 0.01 | 0.08 |
| **FCN2** | 0.05 | 0.36 | 0.72 | 0.96 | -0.07 | -0.39 | 0.70 | 0.93 | -0.06 | -0.58 | 0.56 | 0.85 | 0.24 | 1.91 | 0.06 | 0.30 |
| **FGF2** | -0.12 | -0.33 | 0.74 | 0.96 | 0.33 | 0.87 | 0.39 | 0.74 | 1.16 | 3.29 | 0.00 | 0.03 | 0.03 | 0.09 | 0.93 | 0.93 |
| **FLT1** | -0.29 | -2.32 | 0.02 | 0.28 | -0.26 | -1.80 | 0.08 | 0.34 | -0.12 | -0.88 | 0.38 | 0.67 | 0.02 | 0.16 | 0.87 | 0.92 |
| **FOLR1** | 0.08 | 0.75 | 0.46 | 0.86 | 0.32 | 2.34 | 0.02 | 0.25 | 0.18 | 1.83 | 0.07 | 0.23 | 0.17 | 1.48 | 0.14 | 0.48 |
| **GDF15** | 0.04 | 0.10 | 0.92 | 0.97 | -0.58 | -0.94 | 0.35 | 0.69 | 0.19 | 0.50 | 0.62 | 0.85 | -0.12 | -0.28 | 0.78 | 0.84 |
| **GDI1** | -0.10 | -1.28 | 0.21 | 0.65 | -0.08 | -0.94 | 0.35 | 0.69 | -0.11 | -1.31 | 0.19 | 0.45 | -0.04 | -0.57 | 0.57 | 0.76 |
| **GDNF** | -0.86 | -1.39 | 0.17 | 0.58 | 0.19 | 0.31 | 0.76 | 0.94 | -0.92 | -1.40 | 0.17 | 0.40 | -0.87 | -1.62 | 0.11 | 0.41 |
| **GFAP** | 0.70 | 3.92 | 0.00 | 0.01 | 0.42 | 2.37 | 0.02 | 0.25 | 0.50 | 2.88 | 0.01 | 0.04 | 0.39 | 2.88 | 0.01 | 0.08 |
| **GOT1** | 0.09 | 0.69 | 0.49 | 0.87 | 0.18 | 1.16 | 0.25 | 0.64 | 0.29 | 2.03 | 0.05 | 0.17 | 0.08 | 0.51 | 0.61 | 0.79 |
| **HBA1** | -0.52 | -0.93 | 0.35 | 0.78 | 0.65 | 1.12 | 0.27 | 0.64 | 0.83 | 1.27 | 0.21 | 0.46 | 0.33 | 0.57 | 0.57 | 0.76 |
| **HTT** | -0.08 | -0.32 | 0.75 | 0.96 | 0.45 | 1.44 | 0.16 | 0.53 | 0.71 | 2.24 | 0.03 | 0.14 | 0.30 | 1.11 | 0.27 | 0.58 |
| **ICAM1** | -0.03 | -0.26 | 0.79 | 0.96 | 0.20 | 1.98 | 0.05 | 0.29 | 0.03 | 0.25 | 0.81 | 0.96 | 0.21 | 2.43 | 0.02 | 0.13 |
| **IFNG** | -0.03 | -0.12 | 0.90 | 0.97 | -0.18 | -0.65 | 0.52 | 0.81 | 0.32 | 1.28 | 0.21 | 0.46 | 0.24 | 0.83 | 0.41 | 0.69 |
| **IGF1R** | 0.00 | -0.06 | 0.95 | 0.97 | 0.03 | 0.43 | 0.67 | 0.93 | 0.01 | 0.12 | 0.90 | 0.98 | 0.06 | 1.35 | 0.18 | 0.51 |
| **IGFBP7** | 0.01 | 0.08 | 0.94 | 0.97 | 0.01 | 0.11 | 0.92 | 0.96 | 0.03 | 0.42 | 0.68 | 0.90 | -0.03 | -0.36 | 0.72 | 0.84 |
| **IL10** | -0.12 | -0.33 | 0.74 | 0.96 | -0.15 | -0.47 | 0.64 | 0.90 | -0.38 | -1.36 | 0.18 | 0.43 | 0.04 | 0.12 | 0.90 | 0.92 |
| **IL12p70** | -0.14 | -0.83 | 0.41 | 0.82 | -0.18 | -0.82 | 0.42 | 0.74 | -0.03 | -0.17 | 0.87 | 0.98 | 0.14 | 0.71 | 0.48 | 0.74 |
| **IL13** | -0.11 | -0.34 | 0.74 | 0.96 | 0.08 | 0.23 | 0.82 | 0.94 | 0.06 | 0.22 | 0.82 | 0.96 | 0.12 | 0.41 | 0.68 | 0.84 |
| **IL15** | -0.09 | -0.85 | 0.40 | 0.81 | 0.04 | 0.24 | 0.81 | 0.94 | -0.04 | -0.39 | 0.70 | 0.91 | 0.06 | 0.63 | 0.53 | 0.75 |
| **IL16** | -0.11 | -0.72 | 0.48 | 0.87 | 0.03 | 0.19 | 0.85 | 0.96 | 0.14 | 1.13 | 0.26 | 0.52 | 0.04 | 0.36 | 0.72 | 0.84 |
| **IL17A** | 0.46 | 1.66 | 0.10 | 0.44 | 0.99 | 2.36 | 0.02 | 0.25 | 0.51 | 1.62 | 0.11 | 0.31 | 1.07 | 2.81 | 0.01 | 0.08 |
| **IL18** | 0.11 | 0.62 | 0.54 | 0.88 | 0.41 | 1.91 | 0.06 | 0.30 | 0.74 | 3.12 | 0.00 | 0.04 | 0.30 | 1.69 | 0.10 | 0.41 |
| **IL1B** | 0.07 | 0.98 | 0.33 | 0.77 | 0.16 | 2.02 | 0.05 | 0.29 | 0.29 | 3.15 | 0.00 | 0.04 | 0.13 | 1.96 | 0.05 | 0.29 |
| **IL2** | -0.18 | -1.24 | 0.22 | 0.65 | -0.12 | -0.79 | 0.43 | 0.74 | -0.10 | -0.71 | 0.48 | 0.77 | 0.12 | 0.88 | 0.38 | 0.66 |
| **IL33** | -0.08 | -0.57 | 0.57 | 0.88 | 0.03 | 0.17 | 0.86 | 0.96 | 0.19 | 1.07 | 0.29 | 0.53 | 0.05 | 0.34 | 0.73 | 0.84 |
| **IL4** | -0.48 | -1.61 | 0.11 | 0.46 | -0.05 | -0.16 | 0.88 | 0.96 | 0.53 | 2.03 | 0.05 | 0.17 | 0.08 | 0.28 | 0.78 | 0.84 |
| **IL5** | 0.40 | 1.67 | 0.10 | 0.44 | 0.39 | 1.57 | 0.12 | 0.47 | 0.32 | 1.17 | 0.25 | 0.50 | 0.28 | 1.24 | 0.22 | 0.51 |
| **IL6** | 0.21 | 0.78 | 0.44 | 0.86 | 0.30 | 1.08 | 0.28 | 0.64 | 0.28 | 1.17 | 0.25 | 0.50 | 0.53 | 2.28 | 0.03 | 0.18 |
| **IL6R** | -0.02 | -0.15 | 0.88 | 0.97 | -0.02 | -0.14 | 0.89 | 0.96 | -0.09 | -0.86 | 0.39 | 0.68 | -0.05 | -0.49 | 0.63 | 0.79 |
| **IL7** | -0.09 | -0.56 | 0.57 | 0.88 | -0.19 | -0.97 | 0.34 | 0.69 | -0.16 | -0.95 | 0.34 | 0.62 | 0.42 | 2.90 | 0.01 | 0.08 |
| **IL9** | -0.22 | -1.19 | 0.24 | 0.66 | -0.23 | -0.83 | 0.41 | 0.74 | -0.29 | -1.41 | 0.16 | 0.40 | 0.00 | 0.02 | 0.98 | 0.98 |
| **KDR** | -0.40 | -1.01 | 0.32 | 0.77 | -0.23 | -0.49 | 0.62 | 0.90 | -0.20 | -0.48 | 0.63 | 0.85 | 0.09 | 0.28 | 0.78 | 0.84 |
| **KLK6** | -0.08 | -0.69 | 0.49 | 0.87 | -0.01 | -0.07 | 0.95 | 0.96 | -0.01 | -0.13 | 0.90 | 0.98 | 0.11 | 1.25 | 0.22 | 0.51 |
| **MAPT** | 0.75 | 5.92 | 0.00 | 0.00 | 0.34 | 1.87 | 0.07 | 0.32 | 0.21 | 1.63 | 0.11 | 0.31 | 0.19 | 1.27 | 0.21 | 0.51 |
| **MDH1** | -0.03 | -0.12 | 0.91 | 0.97 | 0.43 | 1.23 | 0.23 | 0.64 | 0.79 | 2.32 | 0.02 | 0.13 | 0.44 | 1.59 | 0.12 | 0.41 |
| **MME** | -0.75 | -1.88 | 0.06 | 0.38 | 0.04 | 0.09 | 0.93 | 0.96 | 0.31 | 0.69 | 0.49 | 0.78 | 0.24 | 0.49 | 0.63 | 0.79 |
| **MSLN** | -0.16 | -0.65 | 0.52 | 0.88 | 0.21 | 0.71 | 0.48 | 0.76 | -0.04 | -0.15 | 0.88 | 0.98 | 0.08 | 0.34 | 0.74 | 0.84 |
| **NEFH** | 1.41 | 2.25 | 0.03 | 0.28 | 0.57 | 0.83 | 0.41 | 0.74 | 1.00 | 1.58 | 0.12 | 0.32 | 1.45 | 2.19 | 0.03 | 0.20 |
| **NEFL ( or NfL)** | 0.40 | 3.68 | 0.00 | 0.01 | 0.37 | 2.63 | 0.01 | 0.22 | 1.15 | 7.49 | 0.00 | 0.00 | 1.00 | 8.11 | 0.00 | 0.00 |
| **NGF** | 0.00 | 0.04 | 0.97 | 0.97 | 0.03 | 0.28 | 0.78 | 0.94 | 0.00 | 0.02 | 0.98 | 0.99 | 0.17 | 0.99 | 0.33 | 0.62 |
| **NPTX1** | -0.19 | -1.41 | 0.16 | 0.58 | 0.20 | 1.51 | 0.14 | 0.48 | -0.02 | -0.14 | 0.89 | 0.98 | -0.08 | -0.70 | 0.49 | 0.74 |
| **NPTX2** | -0.11 | -0.92 | 0.36 | 0.78 | 0.07 | 0.54 | 0.59 | 0.89 | 0.07 | 0.58 | 0.57 | 0.85 | 0.19 | 1.65 | 0.10 | 0.41 |
| **NPTXR** | -0.21 | -1.49 | 0.14 | 0.53 | -0.06 | -0.46 | 0.65 | 0.90 | -0.15 | -1.16 | 0.25 | 0.50 | 0.07 | 0.61 | 0.55 | 0.75 |
| **NPY** | 1.13 | 3.43 | 0.00 | 0.02 | 0.32 | 0.79 | 0.44 | 0.74 | -0.29 | -0.80 | 0.42 | 0.72 | -0.29 | -0.77 | 0.44 | 0.71 |
| **NRGN** | 0.08 | 0.25 | 0.81 | 0.96 | 0.27 | 0.78 | 0.44 | 0.74 | 0.00 | 0.01 | 0.99 | 0.99 | -0.19 | -0.73 | 0.47 | 0.74 |
| **Oligo-SNCA** | -0.18 | -0.31 | 0.76 | 0.96 | 0.79 | 1.31 | 0.19 | 0.58 | 1.50 | 2.06 | 0.04 | 0.17 | 0.29 | 0.57 | 0.57 | 0.76 |
| **PARK7** | 0.14 | 0.69 | 0.49 | 0.87 | 0.55 | 2.25 | 0.03 | 0.25 | 0.93 | 3.51 | 0.00 | 0.02 | 0.39 | 1.32 | 0.19 | 0.51 |
| **PDGFRB** | -0.01 | -0.06 | 0.95 | 0.97 | 0.16 | 0.84 | 0.41 | 0.74 | 0.00 | 0.01 | 0.99 | 0.99 | 0.22 | 1.27 | 0.21 | 0.51 |
| **PDLIM5** | -0.77 | -1.17 | 0.25 | 0.67 | -0.08 | -0.11 | 0.92 | 0.96 | -0.46 | -0.71 | 0.48 | 0.77 | -0.65 | -1.04 | 0.30 | 0.61 |
| **PGF** | -0.15 | -1.48 | 0.14 | 0.53 | -0.03 | -0.27 | 0.79 | 0.94 | -0.05 | -0.54 | 0.59 | 0.85 | 0.02 | 0.30 | 0.76 | 0.84 |
| **PGK1** | 0.22 | 0.59 | 0.56 | 0.88 | 0.78 | 1.96 | 0.05 | 0.29 | 1.40 | 3.16 | 0.00 | 0.04 | 0.47 | 1.31 | 0.19 | 0.51 |
| **POSTN** | 0.09 | 0.61 | 0.54 | 0.88 | 0.28 | 1.52 | 0.13 | 0.48 | 0.04 | 0.26 | 0.79 | 0.96 | 0.69 | 4.84 | 0.00 | 0.00 |
| **PRDX6** | -0.03 | -0.09 | 0.93 | 0.97 | 0.75 | 2.31 | 0.02 | 0.25 | 1.01 | 3.07 | 0.00 | 0.04 | 0.26 | 0.97 | 0.34 | 0.63 |
| **PSEN1** | 0.26 | 1.94 | 0.06 | 0.35 | 0.29 | 2.19 | 0.03 | 0.26 | 0.47 | 3.85 | 0.00 | 0.02 | 0.33 | 3.35 | 0.00 | 0.04 |
| **pSNCA-129** | 0.10 | 0.30 | 0.77 | 0.96 | 0.97 | 2.88 | 0.01 | 0.13 | 1.21 | 3.09 | 0.00 | 0.04 | 0.30 | 0.90 | 0.37 | 0.65 |
| **p-tau-181** | 1.02 | 8.09 | 0.00 | 0.00 | 0.63 | 3.11 | 0.00 | 0.10 | 0.21 | 1.50 | 0.14 | 0.36 | 0.37 | 2.47 | 0.02 | 0.12 |
| **p-tau-217** | 1.07 | 8.40 | 0.00 | 0.00 | 0.55 | 3.27 | 0.00 | 0.10 | 0.12 | 1.05 | 0.30 | 0.54 | 0.10 | 0.87 | 0.39 | 0.67 |
| **p-tau-231** | 1.29 | 9.33 | 0.00 | 0.00 | 0.80 | 3.82 | 0.00 | 0.04 | 0.22 | 1.53 | 0.13 | 0.35 | 0.43 | 2.67 | 0.01 | 0.09 |
| **pTDP43-409** | -0.23 | -1.64 | 0.11 | 0.45 | 0.15 | 0.74 | 0.46 | 0.76 | 0.04 | 0.22 | 0.83 | 0.96 | -0.09 | -0.62 | 0.54 | 0.75 |
| **PTN** | 0.01 | 0.19 | 0.85 | 0.97 | 0.01 | 0.08 | 0.93 | 0.96 | -0.07 | -1.18 | 0.24 | 0.50 | 0.02 | 0.35 | 0.73 | 0.84 |
| **REST** | -0.04 | -0.29 | 0.77 | 0.96 | -0.06 | -0.39 | 0.70 | 0.93 | 0.00 | 0.01 | 0.99 | 0.99 | 0.20 | 1.30 | 0.20 | 0.51 |
| **RUVBL2** | -0.19 | -0.64 | 0.52 | 0.88 | 0.31 | 1.01 | 0.32 | 0.68 | 0.73 | 2.20 | 0.03 | 0.14 | 0.17 | 0.65 | 0.52 | 0.75 |
| **S100A12** | 0.18 | 0.59 | 0.56 | 0.88 | 0.49 | 1.62 | 0.11 | 0.44 | 1.03 | 3.46 | 0.00 | 0.02 | 0.72 | 2.67 | 0.01 | 0.09 |
| **S100B** | 0.12 | 0.99 | 0.33 | 0.77 | 0.05 | 0.37 | 0.71 | 0.93 | -0.01 | -0.08 | 0.93 | 0.99 | 0.28 | 1.60 | 0.11 | 0.41 |
| **SAA1** | 0.11 | 0.24 | 0.81 | 0.96 | 0.92 | 2.14 | 0.04 | 0.26 | 0.24 | 0.59 | 0.56 | 0.85 | 0.83 | 2.04 | 0.05 | 0.26 |
| **SFRP1** | 0.15 | 0.35 | 0.73 | 0.96 | 0.34 | 0.76 | 0.45 | 0.75 | -0.42 | -1.10 | 0.27 | 0.53 | -0.33 | -0.90 | 0.37 | 0.65 |
| **SFTPD** | -0.35 | -1.97 | 0.05 | 0.34 | -0.13 | -0.65 | 0.52 | 0.81 | -0.32 | -1.77 | 0.08 | 0.25 | -0.18 | -1.02 | 0.31 | 0.62 |
| **SLIT2** | -0.05 | -0.55 | 0.59 | 0.89 | 0.01 | 0.06 | 0.95 | 0.96 | 0.00 | -0.04 | 0.97 | 0.99 | 0.09 | 1.00 | 0.32 | 0.62 |
| **SMOC1** | -0.67 | -1.85 | 0.07 | 0.38 | 0.02 | 0.04 | 0.97 | 0.97 | -0.44 | -1.09 | 0.28 | 0.53 | 0.14 | 0.29 | 0.78 | 0.84 |
| **SNAP25** | -0.04 | -0.75 | 0.45 | 0.86 | -0.03 | -0.54 | 0.59 | 0.89 | -0.11 | -2.21 | 0.03 | 0.14 | -0.06 | -1.35 | 0.18 | 0.51 |
| **SNCA** | -0.12 | -0.37 | 0.71 | 0.96 | 0.57 | 1.69 | 0.10 | 0.41 | 0.81 | 2.29 | 0.03 | 0.13 | 0.11 | 0.37 | 0.71 | 0.84 |
| **SNCB** | 0.04 | 0.36 | 0.72 | 0.96 | 0.01 | 0.09 | 0.92 | 0.96 | -0.05 | -0.57 | 0.57 | 0.85 | -0.16 | -1.89 | 0.06 | 0.30 |
| **SOD1** | 0.03 | 0.08 | 0.93 | 0.97 | 0.48 | 1.33 | 0.19 | 0.58 | 1.05 | 2.80 | 0.01 | 0.05 | 0.41 | 1.23 | 0.22 | 0.51 |
| **SQSTM1** | 0.03 | 0.21 | 0.83 | 0.97 | 0.35 | 2.10 | 0.04 | 0.27 | 0.66 | 2.62 | 0.01 | 0.07 | 0.59 | 3.58 | 0.00 | 0.03 |
| **TAFA5** | 0.05 | 0.28 | 0.78 | 0.96 | 0.01 | 0.06 | 0.95 | 0.96 | 0.04 | 0.23 | 0.82 | 0.96 | 0.21 | 1.46 | 0.15 | 0.48 |
| **TARDBP** | 0.02 | 0.11 | 0.92 | 0.97 | 0.38 | 1.35 | 0.18 | 0.58 | 0.84 | 2.78 | 0.01 | 0.05 | 0.31 | 1.20 | 0.23 | 0.52 |
| **TEK** | -0.11 | -1.41 | 0.16 | 0.58 | -0.10 | -0.99 | 0.32 | 0.69 | -0.01 | -0.07 | 0.94 | 0.99 | 0.11 | 1.32 | 0.19 | 0.51 |
| **TIMP3** | 0.46 | 1.26 | 0.21 | 0.65 | 0.17 | 0.41 | 0.68 | 0.93 | 0.04 | 0.11 | 0.91 | 0.98 | 0.27 | 0.79 | 0.43 | 0.70 |
| **TNF** | -0.15 | -1.30 | 0.20 | 0.65 | -0.09 | -0.72 | 0.47 | 0.76 | 0.02 | 0.21 | 0.83 | 0.96 | 0.06 | 0.66 | 0.51 | 0.75 |
| **TREM1** | 0.01 | 0.06 | 0.95 | 0.97 | 0.17 | 1.12 | 0.27 | 0.64 | 0.02 | 0.21 | 0.83 | 0.96 | 0.22 | 1.89 | 0.06 | 0.30 |
| **TREM2** | 0.11 | 0.73 | 0.47 | 0.87 | 0.38 | 2.15 | 0.04 | 0.26 | 0.30 | 2.08 | 0.04 | 0.17 | 0.34 | 2.74 | 0.01 | 0.08 |
| **UCHL1** | -0.07 | -1.14 | 0.26 | 0.69 | -0.01 | -0.13 | 0.90 | 0.96 | -0.10 | -1.75 | 0.09 | 0.26 | 0.01 | 0.12 | 0.91 | 0.92 |
| **VCAM1** | -0.06 | -0.60 | 0.55 | 0.88 | -0.05 | -0.41 | 0.69 | 0.93 | 0.00 | 0.03 | 0.98 | 0.99 | 0.10 | 1.37 | 0.18 | 0.51 |
| **VEGFA** | 0.17 | 0.97 | 0.33 | 0.77 | -0.17 | -0.95 | 0.35 | 0.69 | 0.18 | 1.11 | 0.27 | 0.53 | 0.17 | 0.99 | 0.32 | 0.62 |
| **VEGFD** | -0.11 | -0.97 | 0.34 | 0.77 | 0.22 | 1.39 | 0.17 | 0.56 | 0.04 | 0.40 | 0.69 | 0.91 | 0.02 | 0.15 | 0.88 | 0.92 |
| **VGF** | -1.16 | -2.09 | 0.04 | 0.30 | -1.00 | -1.54 | 0.13 | 0.48 | -1.11 | -1.87 | 0.07 | 0.22 | -0.70 | -1.25 | 0.22 | 0.51 |
| **VSNL1** | -0.02 | -0.22 | 0.83 | 0.97 | 0.18 | 1.18 | 0.24 | 0.64 | 0.09 | 0.79 | 0.43 | 0.72 | 0.18 | 1.67 | 0.10 | 0.41 |
| **YWHAZ** | 0.10 | 1.22 | 0.23 | 0.65 | 0.10 | 0.95 | 0.34 | 0.69 | 0.17 | 2.40 | 0.02 | 0.11 | 0.11 | 1.28 | 0.21 | 0.51 |

Supplementary Table 4. LIMMA full results on NULISA Inflammation panel markers, comparing each diagnostic group with controls. Uncorrected p values < 0.05 are highlighted in yellow, while adjusted p values < 0.05 are highlighted in red.

|  | **Controls vs AD/MCI** | | | | **Controls vs LBD** | | | | **Controls vs FTD** | | | | **Controls vs PSP** | | | |
| --- | --- | --- | --- | --- | --- | --- | --- | --- | --- | --- | --- | --- | --- | --- | --- | --- |
|  | **logFC** | **t** | **p** | **adj p** | **logFC** | **t** | **p** | **adj p** | **logFC** | **t** | **p** | **adj p** | **logFC** | **t** | **p** | **adj p** |
| **AGRP** | -0.06 | -0.65 | 0.52 | 0.94 | 0.15 | 1.71 | 0.09 | 0.46 | 0.09 | 1.12 | 0.27 | 0.57 | 0.12 | 1.56 | 0.12 | 0.32 |
| **ANGPT1** | -0.13 | -0.78 | 0.44 | 0.89 | -0.31 | -1.67 | 0.10 | 0.46 | 0.07 | 0.42 | 0.68 | 0.88 | -0.19 | -1.18 | 0.24 | 0.46 |
| **ANGPT2** | 0.00 | 0.03 | 0.98 | 0.99 | 0.19 | 1.26 | 0.21 | 0.63 | 0.13 | 1.15 | 0.25 | 0.55 | 0.20 | 1.94 | 0.06 | 0.21 |
| **ANXA1** | -0.14 | -0.25 | 0.80 | 0.98 | -0.13 | -0.25 | 0.80 | 0.95 | 0.17 | 0.32 | 0.75 | 0.91 | 0.27 | 0.59 | 0.56 | 0.74 |
| **AREG** | 0.23 | 2.93 | 0.00 | 0.33 | 0.39 | 4.18 | 0.00 | 0.02 | 0.38 | 4.32 | 0.00 | 0.01 | 0.34 | 3.72 | 0.00 | 0.01 |
| **BDNF** | 0.09 | 0.89 | 0.38 | 0.88 | -0.11 | -0.96 | 0.34 | 0.74 | 0.06 | 0.66 | 0.51 | 0.77 | 0.06 | 0.63 | 0.53 | 0.72 |
| **BMP7** | 0.08 | 0.91 | 0.37 | 0.88 | 0.17 | 1.95 | 0.06 | 0.42 | 0.12 | 1.29 | 0.20 | 0.49 | 0.07 | 0.82 | 0.42 | 0.64 |
| **BST2** | -0.01 | -0.03 | 0.97 | 0.99 | 0.14 | 0.79 | 0.43 | 0.83 | 0.21 | 1.15 | 0.25 | 0.55 | 0.11 | 0.70 | 0.48 | 0.67 |
| **C1QA** | 0.01 | 0.06 | 0.95 | 0.99 | 0.25 | 1.55 | 0.13 | 0.50 | 0.09 | 0.67 | 0.50 | 0.77 | 0.18 | 1.48 | 0.15 | 0.33 |
| **CALCA** | -0.13 | -0.75 | 0.45 | 0.91 | -0.10 | -0.62 | 0.54 | 0.85 | 0.12 | 0.73 | 0.47 | 0.75 | 0.40 | 2.46 | 0.02 | 0.09 |
| **CCL1** | 0.10 | 0.82 | 0.42 | 0.89 | 0.03 | 0.18 | 0.86 | 0.98 | 0.02 | 0.11 | 0.92 | 0.99 | 0.13 | 1.10 | 0.27 | 0.50 |
| **CCL11** | 0.31 | 2.51 | 0.01 | 0.45 | 0.23 | 1.77 | 0.08 | 0.46 | 0.25 | 1.88 | 0.07 | 0.24 | 0.18 | 1.53 | 0.13 | 0.32 |
| **CCL13** | 0.37 | 2.28 | 0.03 | 0.53 | 0.19 | 1.26 | 0.21 | 0.63 | 0.33 | 2.58 | 0.01 | 0.09 | 0.12 | 0.90 | 0.37 | 0.60 |
| **CCL14** | 0.08 | 0.92 | 0.36 | 0.87 | 0.01 | 0.07 | 0.95 | 0.99 | 0.08 | 0.99 | 0.33 | 0.63 | 0.11 | 1.46 | 0.15 | 0.34 |
| **CCL15** | 0.07 | 0.34 | 0.73 | 0.97 | 0.05 | 0.24 | 0.81 | 0.95 | 0.22 | 1.28 | 0.20 | 0.49 | 0.30 | 1.67 | 0.10 | 0.29 |
| **CCL16** | 0.05 | 0.32 | 0.75 | 0.97 | 0.23 | 1.83 | 0.07 | 0.44 | 0.31 | 2.13 | 0.04 | 0.17 | 0.23 | 1.75 | 0.09 | 0.26 |
| **CCL17** | 0.54 | 2.06 | 0.04 | 0.54 | 0.15 | 0.50 | 0.62 | 0.89 | 0.12 | 0.43 | 0.67 | 0.88 | -0.18 | -0.68 | 0.50 | 0.69 |
| **CCL19** | 0.02 | 0.07 | 0.95 | 0.99 | 0.12 | 0.39 | 0.70 | 0.92 | 0.27 | 1.02 | 0.31 | 0.61 | 0.04 | 0.20 | 0.84 | 0.93 |
| **CCL2** | 0.00 | 0.01 | 1.00 | 1.00 | -0.03 | -0.24 | 0.81 | 0.95 | 0.36 | 3.37 | 0.00 | 0.02 | 0.05 | 0.51 | 0.61 | 0.79 |
| **CCL20** | 0.37 | 2.42 | 0.02 | 0.50 | 0.40 | 3.04 | 0.00 | 0.12 | 0.30 | 1.45 | 0.15 | 0.43 | 0.14 | 1.17 | 0.25 | 0.46 |
| **CCL21** | -0.09 | -0.72 | 0.47 | 0.92 | 0.16 | 1.14 | 0.26 | 0.67 | 0.05 | 0.37 | 0.71 | 0.91 | 0.05 | 0.44 | 0.66 | 0.81 |
| **CCL22** | -0.43 | -2.23 | 0.03 | 0.53 | -0.43 | -2.10 | 0.04 | 0.35 | -0.27 | -1.35 | 0.18 | 0.47 | -0.17 | -0.91 | 0.37 | 0.60 |
| **CCL23** | -0.01 | -0.12 | 0.90 | 0.99 | -0.09 | -0.80 | 0.43 | 0.83 | 0.01 | 0.08 | 0.93 | 0.99 | -0.09 | -0.72 | 0.48 | 0.67 |
| **CCL24** | -0.20 | -0.71 | 0.48 | 0.92 | 0.20 | 0.67 | 0.51 | 0.84 | -0.23 | -0.83 | 0.41 | 0.70 | 0.21 | 0.81 | 0.42 | 0.64 |
| **CCL25** | 0.41 | 2.19 | 0.03 | 0.53 | 0.55 | 3.11 | 0.00 | 0.12 | 0.32 | 1.80 | 0.08 | 0.27 | -0.08 | -0.47 | 0.64 | 0.80 |
| **CCL26** | -0.61 | -1.84 | 0.07 | 0.56 | -0.31 | -1.04 | 0.30 | 0.69 | 0.18 | 0.67 | 0.50 | 0.77 | -0.07 | -0.27 | 0.79 | 0.90 |
| **CCL27** | -0.01 | -0.09 | 0.93 | 0.99 | 0.06 | 0.37 | 0.71 | 0.94 | 0.19 | 1.20 | 0.23 | 0.53 | 0.10 | 0.54 | 0.59 | 0.77 |
| **CCL28** | 0.27 | 0.70 | 0.49 | 0.92 | 0.55 | 1.99 | 0.05 | 0.40 | 0.16 | 0.55 | 0.58 | 0.83 | -0.13 | -0.48 | 0.63 | 0.80 |
| **CCL3** | -0.42 | -1.98 | 0.05 | 0.56 | 0.01 | 0.03 | 0.97 | 0.99 | 0.22 | 0.90 | 0.37 | 0.66 | -0.07 | -0.37 | 0.71 | 0.83 |
| **CCL4** | -0.17 | -0.71 | 0.48 | 0.92 | 0.06 | 0.22 | 0.83 | 0.96 | -0.08 | -0.32 | 0.75 | 0.91 | -0.06 | -0.26 | 0.79 | 0.90 |
| **CCL5** | 0.34 | 1.12 | 0.27 | 0.84 | -0.15 | -0.51 | 0.61 | 0.88 | 0.36 | 1.30 | 0.20 | 0.49 | 0.36 | 1.35 | 0.18 | 0.39 |
| **CCL7** | 0.43 | 1.89 | 0.06 | 0.56 | 0.43 | 2.77 | 0.01 | 0.17 | 0.65 | 3.54 | 0.00 | 0.02 | 1.11 | 7.17 | 0.00 | 0.00 |
| **CCL8** | 0.59 | 3.10 | 0.00 | 0.33 | 0.06 | 0.43 | 0.67 | 0.90 | 0.21 | 1.51 | 0.13 | 0.40 | 0.18 | 1.20 | 0.23 | 0.46 |
| **CD200** | 2.85 | 2.55 | 0.01 | 0.45 | 3.37 | 2.40 | 0.02 | 0.24 | 2.91 | 2.65 | 0.01 | 0.08 | 1.43 | 1.04 | 0.30 | 0.52 |
| **CD200R1** | -0.18 | -1.69 | 0.10 | 0.63 | -0.08 | -0.57 | 0.57 | 0.86 | -0.03 | -0.34 | 0.74 | 0.91 | 0.03 | 0.29 | 0.77 | 0.89 |
| **CD27** | -0.08 | -0.54 | 0.59 | 0.95 | -0.01 | -0.05 | 0.96 | 0.99 | 0.05 | 0.31 | 0.76 | 0.91 | 0.02 | 0.14 | 0.89 | 0.95 |
| **CD274** | -0.03 | -0.38 | 0.70 | 0.97 | 0.12 | 1.33 | 0.19 | 0.60 | 0.03 | 0.33 | 0.74 | 0.91 | 0.07 | 0.87 | 0.39 | 0.61 |
| **CD276** | 0.27 | 2.64 | 0.01 | 0.43 | 0.40 | 3.83 | 0.00 | 0.04 | 0.33 | 3.44 | 0.00 | 0.02 | 0.59 | 6.34 | 0.00 | 0.00 |
| **CD3E** | 0.07 | 0.27 | 0.79 | 0.97 | -0.02 | -0.08 | 0.93 | 0.99 | 0.63 | 3.14 | 0.00 | 0.03 | 0.26 | 1.32 | 0.19 | 0.40 |
| **CD4** | 0.02 | 0.30 | 0.77 | 0.97 | 0.09 | 0.93 | 0.36 | 0.75 | 0.00 | 0.02 | 0.98 | 1.00 | 0.13 | 1.82 | 0.07 | 0.25 |
| **CD40** | 0.03 | 0.36 | 0.72 | 0.97 | 0.09 | 1.02 | 0.31 | 0.71 | 0.21 | 2.21 | 0.03 | 0.15 | 0.15 | 1.67 | 0.10 | 0.29 |
| **CD40LG** | 0.48 | 2.29 | 0.03 | 0.53 | -0.51 | -1.73 | 0.09 | 0.46 | 0.31 | 1.32 | 0.19 | 0.47 | 0.09 | 0.44 | 0.66 | 0.81 |
| **CD46** | 0.06 | 0.78 | 0.44 | 0.89 | 0.11 | 1.18 | 0.24 | 0.65 | 0.20 | 2.37 | 0.02 | 0.12 | 0.12 | 1.63 | 0.11 | 0.30 |
| **CD70** | -0.22 | -0.99 | 0.33 | 0.87 | -0.07 | -0.23 | 0.82 | 0.95 | -0.27 | -1.22 | 0.23 | 0.53 | -0.44 | -2.03 | 0.05 | 0.19 |
| **CD80** | -0.13 | -0.89 | 0.37 | 0.88 | -0.12 | -0.69 | 0.49 | 0.84 | 0.03 | 0.20 | 0.84 | 0.96 | 0.00 | 0.01 | 0.99 | 0.99 |
| **CD83** | -0.03 | -0.28 | 0.78 | 0.97 | 0.15 | 1.07 | 0.29 | 0.68 | 0.11 | 0.94 | 0.35 | 0.66 | 0.22 | 1.93 | 0.06 | 0.21 |
| **CD93** | 0.06 | 0.59 | 0.56 | 0.94 | 0.16 | 1.33 | 0.19 | 0.60 | -0.04 | -0.47 | 0.64 | 0.87 | -0.01 | -0.08 | 0.94 | 0.96 |
| **CEACAM5** | -0.01 | -0.03 | 0.97 | 0.99 | 0.33 | 1.67 | 0.10 | 0.46 | 0.18 | 0.83 | 0.41 | 0.70 | 0.29 | 1.34 | 0.19 | 0.39 |
| **CHI3L1** | -0.12 | -0.60 | 0.55 | 0.94 | 0.41 | 1.71 | 0.09 | 0.46 | 0.34 | 1.74 | 0.09 | 0.30 | 0.41 | 2.14 | 0.04 | 0.15 |
| **CLEC4A** | -0.02 | -0.15 | 0.88 | 0.99 | -0.06 | -0.51 | 0.61 | 0.88 | -0.04 | -0.44 | 0.66 | 0.88 | -0.27 | -2.52 | 0.01 | 0.08 |
| **CNTF** | 0.87 | 1.64 | 0.11 | 0.65 | 0.71 | 1.55 | 0.13 | 0.50 | 0.65 | 1.60 | 0.12 | 0.37 | 0.69 | 1.44 | 0.15 | 0.35 |
| **CRP** | -0.22 | -0.56 | 0.58 | 0.94 | 0.17 | 0.46 | 0.65 | 0.90 | 0.21 | 0.56 | 0.58 | 0.83 | 0.45 | 1.22 | 0.23 | 0.45 |
| **CSF1** | 0.21 | 2.15 | 0.04 | 0.53 | 0.26 | 2.37 | 0.02 | 0.24 | 0.36 | 3.60 | 0.00 | 0.02 | 0.32 | 3.10 | 0.00 | 0.03 |
| **CSF1R** | -0.01 | -0.09 | 0.93 | 0.99 | 0.25 | 1.30 | 0.20 | 0.62 | 0.10 | 0.62 | 0.54 | 0.79 | 0.32 | 2.25 | 0.03 | 0.13 |
| **CSF2** | -0.18 | -1.56 | 0.12 | 0.70 | -0.06 | -0.41 | 0.68 | 0.91 | -0.02 | -0.16 | 0.87 | 0.98 | -0.01 | -0.06 | 0.96 | 0.97 |
| **CSF2RB** | 0.12 | 0.55 | 0.58 | 0.94 | 0.28 | 1.10 | 0.28 | 0.68 | 0.17 | 0.71 | 0.48 | 0.75 | 0.41 | 1.78 | 0.08 | 0.26 |
| **CSF3** | 0.28 | 1.87 | 0.07 | 0.56 | 0.41 | 2.29 | 0.03 | 0.26 | 0.33 | 2.12 | 0.04 | 0.17 | 0.64 | 4.87 | 0.00 | 0.00 |
| **CSF3R** | 0.00 | 0.01 | 0.99 | 1.00 | 0.15 | 1.36 | 0.18 | 0.60 | 0.10 | 0.92 | 0.36 | 0.66 | 0.25 | 2.73 | 0.01 | 0.06 |
| **CST7** | 0.38 | 1.27 | 0.21 | 0.78 | 0.06 | 0.23 | 0.82 | 0.95 | 0.64 | 1.89 | 0.06 | 0.24 | 0.66 | 1.65 | 0.10 | 0.29 |
| **CTF1** | 0.40 | 1.75 | 0.08 | 0.61 | 0.61 | 2.38 | 0.02 | 0.24 | 0.97 | 3.92 | 0.00 | 0.01 | 0.41 | 1.98 | 0.05 | 0.20 |
| **CTLA4** | -0.11 | -0.80 | 0.43 | 0.89 | 0.04 | 0.24 | 0.81 | 0.95 | 0.14 | 0.90 | 0.37 | 0.66 | 0.04 | 0.32 | 0.75 | 0.87 |
| **CTSS** | -1.86 | -1.11 | 0.27 | 0.84 | -1.16 | -0.62 | 0.54 | 0.85 | 0.58 | 0.35 | 0.73 | 0.91 | 1.04 | 0.64 | 0.53 | 0.72 |
| **CX3CL1** | -0.09 | -0.97 | 0.34 | 0.87 | 0.00 | 0.01 | 0.99 | 0.99 | 0.14 | 1.37 | 0.17 | 0.47 | 0.42 | 3.45 | 0.00 | 0.02 |
| **CXADR** | -0.24 | -2.04 | 0.05 | 0.54 | 0.14 | 0.89 | 0.38 | 0.75 | 0.05 | 0.31 | 0.76 | 0.91 | 0.21 | 1.49 | 0.14 | 0.33 |
| **CXCL1** | -0.20 | -1.05 | 0.30 | 0.86 | -0.35 | -1.67 | 0.10 | 0.46 | 0.30 | 1.17 | 0.25 | 0.55 | -0.09 | -0.48 | 0.63 | 0.80 |
| **CXCL10** | 0.19 | 1.08 | 0.29 | 0.86 | 0.37 | 1.91 | 0.06 | 0.43 | 0.43 | 2.23 | 0.03 | 0.15 | 0.43 | 2.50 | 0.02 | 0.09 |
| **CXCL11** | 0.25 | 0.88 | 0.38 | 0.88 | 0.47 | 1.80 | 0.08 | 0.45 | 0.60 | 2.52 | 0.01 | 0.10 | 0.56 | 2.68 | 0.01 | 0.07 |
| **CXCL12** | -0.17 | -0.27 | 0.79 | 0.97 | -0.91 | -1.30 | 0.20 | 0.62 | -0.74 | -1.09 | 0.28 | 0.58 | -0.05 | -0.08 | 0.94 | 0.96 |
| **CXCL13** | -0.14 | -0.57 | 0.57 | 0.94 | -0.42 | -1.51 | 0.14 | 0.52 | -0.07 | -0.24 | 0.81 | 0.95 | -0.11 | -0.43 | 0.67 | 0.81 |
| **CXCL14** | 0.55 | 1.34 | 0.18 | 0.77 | 0.81 | 1.51 | 0.14 | 0.52 | 0.28 | 0.66 | 0.51 | 0.77 | 0.09 | 0.23 | 0.82 | 0.92 |
| **CXCL16** | -0.04 | -0.36 | 0.72 | 0.97 | -0.10 | -0.91 | 0.37 | 0.75 | 0.01 | 0.10 | 0.92 | 0.99 | 0.01 | 0.10 | 0.92 | 0.95 |
| **CXCL2** | 0.27 | 0.94 | 0.35 | 0.87 | -0.34 | -1.28 | 0.21 | 0.63 | 0.81 | 3.15 | 0.00 | 0.03 | 0.39 | 1.55 | 0.13 | 0.32 |
| **CXCL3** | 0.23 | 1.25 | 0.22 | 0.78 | -0.34 | -1.90 | 0.06 | 0.43 | 0.28 | 1.87 | 0.07 | 0.24 | 0.25 | 1.51 | 0.14 | 0.33 |
| **CXCL5** | 0.07 | 0.34 | 0.74 | 0.97 | -0.16 | -0.65 | 0.52 | 0.84 | 0.11 | 0.51 | 0.61 | 0.85 | 0.18 | 0.93 | 0.36 | 0.59 |
| **CXCL6** | -0.09 | -0.38 | 0.70 | 0.97 | -0.02 | -0.09 | 0.93 | 0.99 | 0.29 | 1.18 | 0.24 | 0.55 | 0.17 | 0.74 | 0.46 | 0.67 |
| **CXCL8** | 0.04 | 0.32 | 0.75 | 0.97 | -0.02 | -0.16 | 0.88 | 0.98 | 0.53 | 3.14 | 0.00 | 0.03 | 0.29 | 2.67 | 0.01 | 0.07 |
| **CXCL9** | 0.14 | 0.60 | 0.55 | 0.94 | 0.09 | 0.41 | 0.68 | 0.91 | 0.08 | 0.32 | 0.75 | 0.91 | 0.17 | 0.76 | 0.45 | 0.66 |
| **EGF** | 0.36 | 1.84 | 0.07 | 0.56 | -0.02 | -0.08 | 0.93 | 0.99 | 0.57 | 3.05 | 0.00 | 0.03 | 0.14 | 0.78 | 0.44 | 0.66 |
| **EPO** | 0.02 | 0.20 | 0.84 | 0.99 | 0.23 | 1.74 | 0.09 | 0.46 | 0.30 | 2.29 | 0.03 | 0.14 | 0.26 | 2.36 | 0.02 | 0.10 |
| **FASLG** | 0.12 | 0.94 | 0.35 | 0.87 | 0.10 | 0.73 | 0.47 | 0.84 | 0.34 | 3.09 | 0.00 | 0.03 | 0.07 | 0.61 | 0.55 | 0.73 |
| **FGF19** | -0.58 | -2.21 | 0.03 | 0.53 | -0.31 | -1.06 | 0.29 | 0.68 | -0.20 | -0.76 | 0.45 | 0.73 | -0.22 | -1.05 | 0.30 | 0.52 |
| **FGF2** | -0.05 | -0.13 | 0.89 | 0.99 | 0.38 | 1.07 | 0.29 | 0.68 | 1.16 | 3.49 | 0.00 | 0.02 | 0.08 | 0.23 | 0.82 | 0.92 |
| **FGF21** | 0.19 | 0.78 | 0.44 | 0.89 | 0.24 | 0.94 | 0.35 | 0.75 | 0.50 | 2.02 | 0.05 | 0.20 | 0.38 | 1.73 | 0.09 | 0.27 |
| **FGF23** | -0.37 | -1.13 | 0.26 | 0.84 | -0.19 | -0.56 | 0.58 | 0.86 | -0.30 | -0.85 | 0.40 | 0.68 | -0.15 | -0.49 | 0.63 | 0.80 |
| **FLT1** | -0.03 | -0.34 | 0.74 | 0.97 | -0.15 | -1.25 | 0.21 | 0.63 | 0.12 | 1.14 | 0.26 | 0.55 | 0.16 | 1.62 | 0.11 | 0.30 |
| **FLT3LG** | 0.08 | 0.67 | 0.50 | 0.94 | 0.01 | 0.08 | 0.94 | 0.99 | 0.05 | 0.38 | 0.71 | 0.91 | -0.06 | -0.38 | 0.71 | 0.83 |
| **FLT4** | -0.03 | -0.24 | 0.81 | 0.98 | 0.10 | 0.89 | 0.38 | 0.75 | 0.11 | 1.11 | 0.27 | 0.57 | 0.32 | 3.21 | 0.00 | 0.03 |
| **FTH1** | -0.04 | -0.28 | 0.78 | 0.97 | -0.10 | -0.45 | 0.65 | 0.90 | -0.16 | -0.86 | 0.39 | 0.68 | -0.17 | -0.92 | 0.36 | 0.59 |
| **FURIN** | -0.06 | -0.49 | 0.63 | 0.95 | 0.03 | 0.25 | 0.80 | 0.95 | 0.26 | 1.99 | 0.05 | 0.21 | 0.29 | 2.57 | 0.01 | 0.08 |
| **GDF15** | -0.14 | -0.27 | 0.79 | 0.97 | -0.57 | -0.90 | 0.37 | 0.75 | -0.02 | -0.04 | 0.97 | 0.99 | -0.12 | -0.26 | 0.80 | 0.90 |
| **GDF2** | -0.18 | -1.70 | 0.09 | 0.63 | 0.01 | 0.09 | 0.93 | 0.99 | -0.06 | -0.54 | 0.59 | 0.83 | 0.14 | 1.35 | 0.18 | 0.39 |
| **GFAP** | 0.71 | 3.86 | 0.00 | 0.07 | 0.41 | 2.21 | 0.03 | 0.29 | 0.54 | 2.96 | 0.00 | 0.04 | 0.36 | 2.46 | 0.02 | 0.09 |
| **GRN** | -0.10 | -1.05 | 0.30 | 0.86 | -0.13 | -1.25 | 0.22 | 0.63 | 0.00 | 0.04 | 0.97 | 0.99 | 0.02 | 0.16 | 0.87 | 0.95 |
| **GZMA** | -0.16 | -1.43 | 0.16 | 0.70 | 0.00 | 0.04 | 0.97 | 0.99 | 0.16 | 1.44 | 0.16 | 0.44 | 0.01 | 0.09 | 0.93 | 0.96 |
| **GZMB** | -0.30 | -1.46 | 0.15 | 0.70 | 0.44 | 1.73 | 0.09 | 0.46 | 0.34 | 1.53 | 0.13 | 0.40 | 0.28 | 1.21 | 0.23 | 0.46 |
| **HAVCR1** | 0.12 | 0.62 | 0.53 | 0.94 | 0.68 | 3.42 | 0.00 | 0.09 | 0.50 | 2.57 | 0.01 | 0.09 | 0.50 | 2.66 | 0.01 | 0.07 |
| **HGF** | -0.18 | -1.00 | 0.32 | 0.87 | 0.16 | 0.90 | 0.37 | 0.75 | 0.56 | 3.32 | 0.00 | 0.03 | 0.55 | 3.69 | 0.00 | 0.01 |
| **HLA-DRA** | -0.01 | -0.05 | 0.96 | 0.99 | 0.16 | 0.81 | 0.42 | 0.82 | 0.46 | 2.48 | 0.02 | 0.10 | 0.17 | 1.06 | 0.29 | 0.52 |
| **ICAM1** | -0.11 | -1.09 | 0.28 | 0.85 | 0.17 | 1.89 | 0.06 | 0.43 | 0.04 | 0.35 | 0.73 | 0.91 | 0.25 | 3.13 | 0.00 | 0.03 |
| **ICOSLG** | -0.14 | -1.58 | 0.12 | 0.70 | -0.07 | -0.64 | 0.52 | 0.84 | 0.05 | 0.46 | 0.64 | 0.87 | 0.09 | 1.18 | 0.24 | 0.46 |
| **IFNA1;IFNA13** | 0.75 | 2.11 | 0.04 | 0.53 | 0.98 | 1.85 | 0.07 | 0.43 | 0.95 | 2.98 | 0.00 | 0.04 | 0.88 | 2.65 | 0.01 | 0.07 |
| **IFNA2** | 0.34 | 2.01 | 0.05 | 0.56 | 0.31 | 1.29 | 0.20 | 0.62 | 0.00 | 0.00 | 1.00 | 1.00 | 0.24 | 1.58 | 0.12 | 0.31 |
| **IFNB1** | -0.44 | -2.11 | 0.04 | 0.53 | 0.10 | 0.33 | 0.74 | 0.95 | -0.01 | -0.05 | 0.96 | 0.99 | -0.24 | -1.00 | 0.32 | 0.55 |
| **IFNG** | 0.03 | 0.13 | 0.90 | 0.99 | -0.12 | -0.43 | 0.67 | 0.90 | 0.49 | 1.90 | 0.06 | 0.23 | 0.30 | 1.03 | 0.31 | 0.53 |
| **IFNL1** | 0.01 | 0.03 | 0.97 | 0.99 | 0.18 | 0.75 | 0.45 | 0.84 | 0.30 | 1.35 | 0.18 | 0.47 | -0.05 | -0.18 | 0.86 | 0.94 |
| **IFNL2;IFNL3** | 0.07 | 0.33 | 0.74 | 0.97 | -0.02 | -0.06 | 0.95 | 0.99 | 0.00 | 0.00 | 1.00 | 1.00 | -0.02 | -0.10 | 0.92 | 0.95 |
| **IFNW1** | -0.06 | -0.25 | 0.81 | 0.98 | 0.02 | 0.05 | 0.96 | 0.99 | -0.03 | -0.13 | 0.90 | 0.98 | -0.07 | -0.20 | 0.84 | 0.93 |
| **IKBKG** | 0.27 | 1.46 | 0.15 | 0.70 | 0.43 | 1.82 | 0.07 | 0.44 | 0.78 | 3.31 | 0.00 | 0.03 | 0.42 | 2.21 | 0.03 | 0.14 |
| **IL10** | 0.01 | 0.03 | 0.98 | 0.99 | -0.04 | -0.12 | 0.90 | 0.99 | -0.20 | -0.70 | 0.48 | 0.75 | 0.14 | 0.42 | 0.68 | 0.82 |
| **IL10RB** | -0.08 | -0.87 | 0.39 | 0.88 | 0.10 | 0.97 | 0.34 | 0.74 | 0.07 | 0.79 | 0.43 | 0.72 | 0.11 | 1.55 | 0.13 | 0.32 |
| **IL11** | -0.15 | -0.31 | 0.76 | 0.97 | 0.34 | 0.55 | 0.59 | 0.87 | -0.10 | -0.22 | 0.83 | 0.96 | -0.21 | -0.53 | 0.60 | 0.77 |
| **IL12B** | 0.02 | 0.11 | 0.91 | 0.99 | 0.25 | 1.17 | 0.25 | 0.65 | 0.42 | 2.14 | 0.04 | 0.17 | 0.28 | 1.50 | 0.14 | 0.33 |
| **IL12p70** | -0.10 | -0.60 | 0.55 | 0.94 | -0.15 | -0.72 | 0.47 | 0.84 | 0.07 | 0.40 | 0.69 | 0.90 | 0.15 | 0.77 | 0.44 | 0.66 |
| **IL12RB1** | -0.17 | -1.27 | 0.21 | 0.78 | -0.02 | -0.15 | 0.88 | 0.98 | 0.02 | 0.13 | 0.90 | 0.98 | -0.10 | -0.71 | 0.48 | 0.67 |
| **IL13** | -0.21 | -0.51 | 0.61 | 0.95 | 0.10 | 0.26 | 0.80 | 0.95 | 0.06 | 0.19 | 0.85 | 0.96 | 0.16 | 0.45 | 0.66 | 0.81 |
| **IL13RA2** | -0.09 | -0.92 | 0.36 | 0.87 | -0.01 | -0.06 | 0.95 | 0.99 | 0.02 | 0.18 | 0.86 | 0.96 | 0.00 | 0.02 | 0.99 | 0.99 |
| **IL15** | 0.06 | 0.64 | 0.53 | 0.94 | 0.04 | 0.34 | 0.74 | 0.95 | 0.10 | 1.02 | 0.31 | 0.61 | 0.14 | 1.51 | 0.14 | 0.33 |
| **IL15RA** | 0.13 | 1.16 | 0.25 | 0.83 | 0.20 | 1.58 | 0.12 | 0.50 | 0.17 | 1.38 | 0.17 | 0.47 | 0.34 | 2.89 | 0.01 | 0.04 |
| **IL16** | -0.06 | -0.44 | 0.66 | 0.97 | 0.08 | 0.68 | 0.50 | 0.84 | 0.21 | 1.85 | 0.07 | 0.25 | 0.08 | 0.78 | 0.44 | 0.66 |
| **IL17A** | 0.54 | 1.65 | 0.10 | 0.65 | 1.10 | 2.44 | 0.02 | 0.24 | 0.74 | 2.00 | 0.05 | 0.21 | 1.27 | 2.96 | 0.00 | 0.04 |
| **IL17A-IL17F** | 0.73 | 2.67 | 0.01 | 0.43 | 0.96 | 3.02 | 0.00 | 0.12 | 0.32 | 1.24 | 0.22 | 0.51 | 0.72 | 3.03 | 0.00 | 0.03 |
| **IL17B** | 0.09 | 0.39 | 0.70 | 0.97 | 0.23 | 1.06 | 0.29 | 0.68 | 0.01 | 0.06 | 0.96 | 0.99 | 0.26 | 1.08 | 0.28 | 0.51 |
| **IL17C** | 0.36 | 1.44 | 0.15 | 0.70 | 0.40 | 1.88 | 0.06 | 0.43 | 0.33 | 1.35 | 0.18 | 0.47 | 0.36 | 1.56 | 0.12 | 0.32 |
| **IL17F** | 0.88 | 1.36 | 0.18 | 0.75 | 0.17 | 0.25 | 0.80 | 0.95 | 0.90 | 1.56 | 0.12 | 0.38 | 0.58 | 0.95 | 0.35 | 0.58 |
| **IL17RA** | -0.03 | -0.41 | 0.68 | 0.97 | 0.02 | 0.27 | 0.79 | 0.95 | 0.02 | 0.35 | 0.73 | 0.91 | 0.10 | 1.76 | 0.08 | 0.26 |
| **IL17RB** | -0.01 | -0.05 | 0.96 | 0.99 | 0.02 | 0.07 | 0.94 | 0.99 | 0.12 | 0.53 | 0.59 | 0.83 | -0.21 | -1.04 | 0.30 | 0.52 |
| **IL18** | 0.19 | 0.96 | 0.34 | 0.87 | 0.51 | 2.29 | 0.03 | 0.26 | 0.84 | 3.26 | 0.00 | 0.03 | 0.36 | 1.78 | 0.08 | 0.26 |
| **IL18BP** | 0.05 | 0.49 | 0.63 | 0.95 | 0.15 | 1.16 | 0.25 | 0.65 | 0.19 | 1.71 | 0.09 | 0.31 | 0.19 | 2.12 | 0.04 | 0.16 |
| **IL18R1** | -0.03 | -0.32 | 0.75 | 0.97 | 0.17 | 1.54 | 0.13 | 0.50 | 0.27 | 2.59 | 0.01 | 0.09 | 0.27 | 3.17 | 0.00 | 0.03 |
| **IL19** | -0.01 | -0.06 | 0.96 | 0.99 | 0.39 | 1.70 | 0.09 | 0.46 | 0.01 | 0.06 | 0.95 | 0.99 | 0.32 | 1.86 | 0.07 | 0.23 |
| **IL1B** | 0.10 | 1.28 | 0.21 | 0.78 | 0.13 | 1.71 | 0.09 | 0.46 | 0.42 | 3.76 | 0.00 | 0.01 | 0.15 | 1.81 | 0.07 | 0.25 |
| **IL1R1** | -0.07 | -0.80 | 0.42 | 0.89 | 0.08 | 0.66 | 0.51 | 0.84 | -0.02 | -0.25 | 0.81 | 0.95 | -0.20 | -1.72 | 0.09 | 0.27 |
| **IL1R2** | -0.04 | -0.50 | 0.62 | 0.95 | 0.05 | 0.47 | 0.64 | 0.90 | -0.02 | -0.24 | 0.81 | 0.95 | 0.05 | 0.56 | 0.58 | 0.76 |
| **IL1RL1** | -0.24 | -1.86 | 0.07 | 0.56 | 0.09 | 0.70 | 0.49 | 0.84 | 0.14 | 1.05 | 0.30 | 0.60 | 0.02 | 0.15 | 0.88 | 0.95 |
| **IL1RN** | 0.14 | 0.85 | 0.40 | 0.89 | 0.23 | 1.55 | 0.13 | 0.50 | 0.67 | 4.17 | 0.00 | 0.01 | 0.76 | 4.66 | 0.00 | 0.00 |
| **IL2** | -0.19 | -1.21 | 0.23 | 0.79 | -0.09 | -0.57 | 0.57 | 0.86 | -0.01 | -0.04 | 0.97 | 0.99 | 0.19 | 1.39 | 0.17 | 0.38 |
| **IL20** | -0.01 | -0.10 | 0.92 | 0.99 | 0.22 | 1.39 | 0.17 | 0.57 | 0.16 | 1.01 | 0.31 | 0.61 | 0.24 | 1.83 | 0.07 | 0.25 |
| **IL22** | 0.52 | 1.75 | 0.09 | 0.61 | 0.60 | 2.01 | 0.05 | 0.39 | 0.69 | 2.44 | 0.02 | 0.11 | 0.77 | 2.65 | 0.01 | 0.07 |
| **IL23** | 0.35 | 1.55 | 0.13 | 0.70 | 0.33 | 1.40 | 0.17 | 0.57 | 0.35 | 1.46 | 0.15 | 0.43 | 0.46 | 2.41 | 0.02 | 0.10 |
| **IL24** | -0.03 | -0.06 | 0.95 | 0.99 | -0.62 | -1.13 | 0.26 | 0.67 | -0.41 | -0.87 | 0.39 | 0.68 | 0.38 | 0.80 | 0.43 | 0.65 |
| **IL27** | -0.46 | -1.37 | 0.17 | 0.75 | 0.03 | 0.09 | 0.93 | 0.99 | 0.01 | 0.03 | 0.97 | 0.99 | 0.25 | 0.73 | 0.47 | 0.67 |
| **IL2RA** | 0.03 | 0.20 | 0.84 | 0.99 | -0.08 | -0.45 | 0.66 | 0.90 | 0.13 | 0.72 | 0.48 | 0.75 | 0.13 | 0.76 | 0.45 | 0.66 |
| **IL2RB** | -0.02 | -0.17 | 0.86 | 0.99 | 0.06 | 0.41 | 0.68 | 0.91 | 0.10 | 0.91 | 0.37 | 0.66 | 0.13 | 1.08 | 0.28 | 0.51 |
| **IL32** | -0.31 | -0.66 | 0.51 | 0.94 | -0.22 | -0.46 | 0.65 | 0.90 | -0.70 | -1.63 | 0.11 | 0.35 | -0.28 | -0.66 | 0.51 | 0.71 |
| **IL33** | 0.04 | 0.30 | 0.77 | 0.97 | 0.14 | 0.77 | 0.44 | 0.84 | 0.36 | 2.27 | 0.03 | 0.14 | 0.20 | 1.58 | 0.12 | 0.31 |
| **IL34** | -0.13 | -0.42 | 0.67 | 0.97 | -0.01 | -0.02 | 0.98 | 0.99 | 0.05 | 0.13 | 0.90 | 0.98 | 0.17 | 0.48 | 0.63 | 0.80 |
| **IL36A** | -0.04 | -0.19 | 0.85 | 0.99 | 0.13 | 0.67 | 0.50 | 0.84 | 0.30 | 1.67 | 0.10 | 0.33 | 0.27 | 1.71 | 0.09 | 0.28 |
| **IL36B** | 0.02 | 0.11 | 0.91 | 0.99 | 0.01 | 0.08 | 0.94 | 0.99 | 0.41 | 2.26 | 0.03 | 0.14 | 0.06 | 0.40 | 0.69 | 0.82 |
| **IL36G** | -0.02 | -0.18 | 0.86 | 0.99 | 0.01 | 0.13 | 0.89 | 0.99 | -0.03 | -0.44 | 0.66 | 0.88 | 0.26 | 2.37 | 0.02 | 0.10 |
| **IL3RA** | -0.10 | -0.82 | 0.42 | 0.89 | 0.18 | 1.45 | 0.15 | 0.55 | -0.17 | -1.47 | 0.15 | 0.43 | 0.05 | 0.42 | 0.67 | 0.82 |
| **IL4** | -0.46 | -1.43 | 0.16 | 0.70 | 0.09 | 0.27 | 0.79 | 0.95 | 0.66 | 2.30 | 0.02 | 0.14 | 0.22 | 0.73 | 0.47 | 0.67 |
| **IL4R** | -0.07 | -0.88 | 0.38 | 0.88 | -0.04 | -0.45 | 0.66 | 0.90 | 0.01 | 0.12 | 0.90 | 0.98 | 0.11 | 1.61 | 0.11 | 0.30 |
| **IL5** | 0.29 | 1.14 | 0.26 | 0.84 | 0.25 | 0.94 | 0.35 | 0.74 | 0.26 | 0.92 | 0.36 | 0.66 | 0.19 | 0.84 | 0.41 | 0.64 |
| **IL5RA** | -0.12 | -0.56 | 0.58 | 0.94 | -0.08 | -0.32 | 0.75 | 0.95 | -0.01 | -0.05 | 0.96 | 0.99 | 0.24 | 1.15 | 0.25 | 0.47 |
| **IL6** | 0.26 | 0.95 | 0.35 | 0.87 | 0.31 | 1.10 | 0.28 | 0.68 | 0.38 | 1.54 | 0.13 | 0.40 | 0.55 | 2.32 | 0.02 | 0.11 |
| **IL6R** | -0.01 | -0.12 | 0.91 | 0.99 | 0.02 | 0.21 | 0.84 | 0.96 | 0.00 | -0.01 | 0.99 | 1.00 | 0.01 | 0.10 | 0.92 | 0.95 |
| **IL6ST** | -0.09 | -1.28 | 0.20 | 0.78 | 0.08 | 1.09 | 0.28 | 0.68 | 0.07 | 0.90 | 0.37 | 0.66 | 0.14 | 2.43 | 0.02 | 0.09 |
| **IL7** | -0.04 | -0.23 | 0.82 | 0.99 | -0.13 | -0.66 | 0.51 | 0.84 | -0.06 | -0.34 | 0.74 | 0.91 | 0.47 | 3.05 | 0.00 | 0.03 |
| **IL7R** | -0.27 | -1.81 | 0.08 | 0.59 | -0.24 | -1.57 | 0.12 | 0.50 | -0.25 | -1.61 | 0.11 | 0.36 | -0.33 | -2.39 | 0.02 | 0.10 |
| **IL9** | -0.16 | -0.79 | 0.43 | 0.89 | -0.15 | -0.51 | 0.61 | 0.88 | -0.16 | -0.75 | 0.46 | 0.74 | 0.10 | 0.44 | 0.66 | 0.81 |
| **IRAK4** | 0.16 | 0.52 | 0.61 | 0.95 | 0.81 | 2.24 | 0.03 | 0.28 | 1.22 | 3.09 | 0.00 | 0.03 | 0.45 | 1.37 | 0.17 | 0.39 |
| **KDR** | -0.38 | -1.03 | 0.31 | 0.86 | -0.30 | -0.65 | 0.52 | 0.84 | -0.15 | -0.37 | 0.71 | 0.91 | 0.04 | 0.12 | 0.90 | 0.95 |
| **KITLG** | 0.05 | 0.57 | 0.57 | 0.94 | 0.26 | 2.68 | 0.01 | 0.19 | 0.08 | 0.93 | 0.35 | 0.66 | 0.05 | 0.58 | 0.56 | 0.74 |
| **KLRK1** | 0.07 | 0.60 | 0.55 | 0.94 | 0.08 | 0.60 | 0.55 | 0.86 | 0.07 | 0.63 | 0.53 | 0.78 | 0.10 | 0.97 | 0.34 | 0.57 |
| **KNG1** | -0.04 | -0.49 | 0.63 | 0.95 | -0.07 | -0.75 | 0.45 | 0.84 | -0.05 | -0.54 | 0.59 | 0.83 | 0.02 | 0.27 | 0.79 | 0.90 |
| **LAG3** | -0.02 | -0.13 | 0.89 | 0.99 | -0.04 | -0.25 | 0.80 | 0.95 | 0.01 | 0.04 | 0.97 | 0.99 | 0.01 | 0.10 | 0.92 | 0.95 |
| **LAMP3** | 0.01 | 0.09 | 0.93 | 0.99 | 0.56 | 3.03 | 0.00 | 0.12 | 0.16 | 1.01 | 0.32 | 0.61 | 0.18 | 1.18 | 0.24 | 0.46 |
| **LCN2** | 0.10 | 0.57 | 0.57 | 0.94 | 0.25 | 1.58 | 0.12 | 0.50 | 0.64 | 3.78 | 0.00 | 0.01 | 0.27 | 1.70 | 0.09 | 0.28 |
| **LGALS9** | -0.03 | -0.28 | 0.78 | 0.97 | 0.12 | 1.22 | 0.23 | 0.64 | 0.17 | 1.75 | 0.09 | 0.30 | 0.32 | 3.17 | 0.00 | 0.03 |
| **LIF** | 0.09 | 0.62 | 0.54 | 0.94 | 0.33 | 2.40 | 0.02 | 0.24 | 0.41 | 2.66 | 0.01 | 0.08 | 0.35 | 3.02 | 0.00 | 0.03 |
| **LILRB2** | -0.03 | -0.27 | 0.78 | 0.97 | -0.13 | -1.34 | 0.18 | 0.60 | 0.03 | 0.32 | 0.75 | 0.91 | 0.01 | 0.14 | 0.89 | 0.95 |
| **LTA** | -0.10 | -0.83 | 0.41 | 0.89 | -0.05 | -0.24 | 0.81 | 0.95 | -0.11 | -1.20 | 0.24 | 0.53 | -0.14 | -1.39 | 0.17 | 0.38 |
| **LTA-LTB** | -0.69 | -1.23 | 0.22 | 0.79 | -0.47 | -0.82 | 0.41 | 0.81 | 0.60 | 1.07 | 0.29 | 0.59 | -0.31 | -0.60 | 0.55 | 0.73 |
| **MERTK** | -0.12 | -1.25 | 0.22 | 0.78 | -0.06 | -0.67 | 0.50 | 0.84 | -0.01 | -0.07 | 0.94 | 0.99 | 0.00 | 0.01 | 0.99 | 0.99 |
| **MICA** | 0.79 | 1.79 | 0.08 | 0.59 | 0.64 | 1.14 | 0.26 | 0.67 | 0.72 | 1.52 | 0.13 | 0.40 | -0.07 | -0.14 | 0.89 | 0.95 |
| **MICB** | -0.26 | -1.43 | 0.16 | 0.70 | -0.02 | -0.07 | 0.95 | 0.99 | -0.08 | -0.42 | 0.68 | 0.88 | 0.13 | 0.74 | 0.46 | 0.67 |
| **MIF** | 0.09 | 0.28 | 0.78 | 0.97 | 0.81 | 2.35 | 0.02 | 0.24 | 1.25 | 3.24 | 0.00 | 0.03 | 0.51 | 1.53 | 0.13 | 0.32 |
| **MMP1** | 0.10 | 0.42 | 0.67 | 0.97 | 0.08 | 0.30 | 0.77 | 0.95 | 0.22 | 0.95 | 0.34 | 0.65 | 0.26 | 1.05 | 0.30 | 0.52 |
| **MMP12** | 0.15 | 0.94 | 0.35 | 0.87 | 0.16 | 0.98 | 0.33 | 0.73 | 0.08 | 0.48 | 0.63 | 0.86 | 0.08 | 0.54 | 0.59 | 0.77 |
| **MMP3** | 0.25 | 1.67 | 0.10 | 0.63 | 0.05 | 0.29 | 0.77 | 0.95 | 0.17 | 1.10 | 0.28 | 0.57 | 0.24 | 1.64 | 0.11 | 0.30 |
| **MMP8** | -0.27 | -0.84 | 0.41 | 0.89 | 0.09 | 0.26 | 0.79 | 0.95 | 0.99 | 2.90 | 0.01 | 0.04 | 0.64 | 2.11 | 0.04 | 0.16 |
| **MMP9** | -0.15 | -0.51 | 0.61 | 0.95 | 0.27 | 0.91 | 0.37 | 0.75 | 0.98 | 3.53 | 0.00 | 0.02 | 0.85 | 3.43 | 0.00 | 0.02 |
| **MPO** | -0.04 | -0.17 | 0.87 | 0.99 | 0.15 | 0.59 | 0.56 | 0.86 | 0.86 | 4.34 | 0.00 | 0.01 | 0.49 | 2.55 | 0.01 | 0.08 |
| **MUC16** | -0.03 | -0.17 | 0.86 | 0.99 | 0.05 | 0.29 | 0.78 | 0.95 | 0.00 | -0.03 | 0.98 | 0.99 | 0.47 | 3.13 | 0.00 | 0.03 |
| **NAMPT** | 0.22 | 0.71 | 0.48 | 0.92 | 0.23 | 0.74 | 0.46 | 0.84 | 0.88 | 2.81 | 0.01 | 0.06 | 0.25 | 0.76 | 0.45 | 0.66 |
| **NCR1** | 0.07 | 0.63 | 0.53 | 0.94 | 0.15 | 1.17 | 0.25 | 0.65 | 0.20 | 2.02 | 0.05 | 0.20 | 0.13 | 1.18 | 0.24 | 0.46 |
| **NGF** | 0.05 | 0.47 | 0.64 | 0.96 | 0.07 | 0.65 | 0.52 | 0.84 | 0.15 | 1.58 | 0.12 | 0.38 | 0.18 | 0.97 | 0.34 | 0.57 |
| **NTF3** | -0.36 | -0.81 | 0.42 | 0.89 | 0.00 | 0.00 | 1.00 | 1.00 | -0.56 | -1.35 | 0.18 | 0.47 | 0.42 | 0.82 | 0.42 | 0.64 |
| **OSM** | 0.11 | 0.46 | 0.65 | 0.96 | 0.44 | 1.86 | 0.07 | 0.43 | 1.01 | 4.15 | 0.00 | 0.01 | 0.79 | 4.17 | 0.00 | 0.00 |
| **OSMR** | -0.03 | -0.44 | 0.66 | 0.97 | 0.05 | 0.56 | 0.58 | 0.86 | -0.01 | -0.12 | 0.90 | 0.98 | 0.24 | 3.88 | 0.00 | 0.01 |
| **PDCD1** | -0.13 | -0.77 | 0.45 | 0.89 | -0.03 | -0.17 | 0.87 | 0.98 | 0.12 | 0.64 | 0.53 | 0.78 | 0.15 | 0.90 | 0.37 | 0.60 |
| **PDCD1LG2** | 0.05 | 0.62 | 0.54 | 0.94 | 0.28 | 2.96 | 0.00 | 0.12 | 0.21 | 2.32 | 0.02 | 0.13 | 0.25 | 3.59 | 0.00 | 0.02 |
| **PDGFA** | 0.06 | 0.73 | 0.47 | 0.92 | 0.01 | 0.11 | 0.91 | 0.99 | -0.02 | -0.19 | 0.85 | 0.96 | 0.01 | 0.12 | 0.91 | 0.95 |
| **PDGFB** | -0.08 | -0.49 | 0.62 | 0.95 | 0.02 | 0.12 | 0.91 | 0.99 | 0.16 | 1.16 | 0.25 | 0.55 | 0.26 | 1.98 | 0.05 | 0.20 |
| **PGF** | -0.04 | -0.55 | 0.59 | 0.94 | 0.05 | 0.65 | 0.52 | 0.84 | 0.06 | 0.78 | 0.44 | 0.72 | 0.07 | 1.18 | 0.24 | 0.46 |
| **PTX3** | -0.08 | -0.47 | 0.64 | 0.96 | -0.09 | -0.52 | 0.61 | 0.88 | 0.31 | 1.78 | 0.08 | 0.28 | 0.51 | 2.80 | 0.01 | 0.05 |
| **S100A12** | 0.22 | 0.65 | 0.52 | 0.94 | 0.46 | 1.39 | 0.17 | 0.57 | 1.06 | 3.25 | 0.00 | 0.03 | 0.79 | 2.55 | 0.01 | 0.08 |
| **S100A9** | 0.26 | 1.16 | 0.25 | 0.83 | 0.47 | 2.44 | 0.02 | 0.24 | 0.51 | 3.29 | 0.00 | 0.03 | 0.51 | 3.30 | 0.00 | 0.02 |
| **SCG2** | 0.05 | 0.29 | 0.77 | 0.97 | 0.11 | 0.48 | 0.63 | 0.90 | -0.06 | -0.32 | 0.75 | 0.91 | -0.18 | -1.08 | 0.28 | 0.51 |
| **SDC1** | 0.06 | 0.62 | 0.54 | 0.94 | 0.30 | 2.59 | 0.01 | 0.21 | 0.13 | 1.33 | 0.19 | 0.47 | 0.33 | 3.75 | 0.00 | 0.01 |
| **SELE** | -0.07 | -0.42 | 0.67 | 0.97 | 0.12 | 0.78 | 0.44 | 0.83 | 0.03 | 0.20 | 0.85 | 0.96 | 0.13 | 0.89 | 0.38 | 0.60 |
| **SELP** | 0.05 | 0.40 | 0.69 | 0.97 | 0.00 | 0.03 | 0.98 | 0.99 | 0.29 | 2.09 | 0.04 | 0.18 | 0.20 | 1.77 | 0.08 | 0.26 |
| **SIRPA** | 0.18 | 1.47 | 0.15 | 0.70 | 0.37 | 3.10 | 0.00 | 0.12 | 0.16 | 1.36 | 0.18 | 0.47 | 0.26 | 2.52 | 0.01 | 0.08 |
| **SLAMF1** | -0.10 | -0.67 | 0.50 | 0.94 | 0.24 | 1.41 | 0.16 | 0.57 | 0.12 | 0.91 | 0.36 | 0.66 | 0.15 | 1.24 | 0.22 | 0.45 |
| **SPP1** | 0.25 | 1.52 | 0.13 | 0.70 | 0.50 | 2.51 | 0.01 | 0.24 | 0.27 | 1.95 | 0.06 | 0.22 | 0.49 | 3.56 | 0.00 | 0.02 |
| **TAFA5** | 0.07 | 0.39 | 0.70 | 0.97 | 0.03 | 0.20 | 0.84 | 0.97 | 0.09 | 0.47 | 0.64 | 0.87 | 0.28 | 1.93 | 0.06 | 0.21 |
| **TEK** | -0.15 | -1.95 | 0.06 | 0.56 | -0.07 | -0.74 | 0.46 | 0.84 | 0.04 | 0.53 | 0.60 | 0.83 | 0.10 | 1.35 | 0.18 | 0.39 |
| **TGFB1** | 0.05 | 0.51 | 0.61 | 0.95 | 0.00 | 0.01 | 0.99 | 0.99 | 0.12 | 1.41 | 0.16 | 0.46 | 0.02 | 0.27 | 0.78 | 0.90 |
| **TGFB3** | -0.28 | -1.03 | 0.31 | 0.86 | 0.21 | 0.54 | 0.59 | 0.87 | -0.15 | -0.50 | 0.62 | 0.85 | -0.06 | -0.19 | 0.85 | 0.94 |
| **THBS2** | -0.20 | -2.10 | 0.04 | 0.53 | -0.09 | -0.72 | 0.47 | 0.84 | 0.02 | 0.20 | 0.85 | 0.96 | 0.04 | 0.39 | 0.70 | 0.83 |
| **THPO** | -0.08 | -0.97 | 0.34 | 0.87 | -0.06 | -0.62 | 0.54 | 0.85 | 0.17 | 1.97 | 0.05 | 0.21 | 0.17 | 2.02 | 0.05 | 0.19 |
| **TIMP1** | -0.01 | -0.04 | 0.97 | 0.99 | -0.20 | -1.22 | 0.23 | 0.64 | -0.25 | -1.44 | 0.15 | 0.44 | -0.06 | -0.40 | 0.69 | 0.82 |
| **TIMP2** | 0.09 | 1.26 | 0.21 | 0.78 | 0.09 | 1.06 | 0.29 | 0.68 | 0.14 | 1.99 | 0.05 | 0.21 | 0.24 | 3.50 | 0.00 | 0.02 |
| **TLR3** | -0.20 | -0.98 | 0.33 | 0.87 | -0.06 | -0.25 | 0.80 | 0.95 | 0.13 | 0.66 | 0.51 | 0.77 | -0.21 | -1.11 | 0.27 | 0.50 |
| **TNF** | -0.14 | -1.21 | 0.23 | 0.79 | -0.03 | -0.26 | 0.79 | 0.95 | 0.11 | 1.00 | 0.32 | 0.62 | 0.12 | 1.24 | 0.22 | 0.45 |
| **TNFRSF11A** | 0.02 | 0.18 | 0.86 | 0.99 | 0.19 | 1.48 | 0.14 | 0.53 | 0.09 | 0.78 | 0.44 | 0.72 | 0.29 | 3.06 | 0.00 | 0.03 |
| **TNFRSF11B** | -0.15 | -1.91 | 0.06 | 0.56 | -0.15 | -1.66 | 0.10 | 0.46 | 0.02 | 0.19 | 0.85 | 0.96 | 0.01 | 0.13 | 0.90 | 0.95 |
| **TNFRSF13B** | -0.32 | -1.38 | 0.17 | 0.75 | -0.27 | -1.00 | 0.32 | 0.73 | -0.06 | -0.24 | 0.81 | 0.95 | -0.23 | -1.00 | 0.32 | 0.55 |
| **TNFRSF13C** | -0.22 | -1.07 | 0.29 | 0.86 | -0.34 | -1.40 | 0.17 | 0.57 | 0.18 | 0.85 | 0.40 | 0.68 | -0.09 | -0.42 | 0.68 | 0.82 |
| **TNFRSF14** | 0.19 | 2.09 | 0.04 | 0.53 | 0.08 | 0.75 | 0.46 | 0.84 | 0.26 | 2.89 | 0.01 | 0.04 | 0.17 | 2.21 | 0.03 | 0.14 |
| **TNFRSF17** | -0.16 | -1.07 | 0.29 | 0.86 | 0.05 | 0.23 | 0.82 | 0.95 | -0.04 | -0.29 | 0.77 | 0.92 | 0.03 | 0.24 | 0.81 | 0.91 |
| **TNFRSF18** | -0.03 | -0.31 | 0.76 | 0.97 | 0.09 | 0.97 | 0.34 | 0.74 | -0.01 | -0.15 | 0.88 | 0.98 | 0.18 | 1.93 | 0.06 | 0.21 |
| **TNFRSF1A** | 0.12 | 0.98 | 0.33 | 0.87 | 0.16 | 1.18 | 0.24 | 0.65 | 0.28 | 2.48 | 0.02 | 0.10 | 0.37 | 3.54 | 0.00 | 0.02 |
| **TNFRSF1B** | -0.08 | -0.56 | 0.58 | 0.94 | 0.10 | 0.66 | 0.51 | 0.84 | 0.08 | 0.61 | 0.55 | 0.80 | 0.26 | 2.18 | 0.03 | 0.14 |
| **TNFRSF21** | 0.01 | 0.05 | 0.96 | 0.99 | 0.13 | 0.95 | 0.34 | 0.74 | 0.06 | 0.57 | 0.57 | 0.83 | 0.01 | 0.07 | 0.94 | 0.96 |
| **TNFRSF4** | -0.17 | -1.15 | 0.25 | 0.83 | -0.03 | -0.18 | 0.86 | 0.98 | -0.02 | -0.14 | 0.89 | 0.98 | 0.09 | 0.61 | 0.55 | 0.73 |
| **TNFRSF8** | -0.14 | -0.98 | 0.33 | 0.87 | -0.02 | -0.17 | 0.86 | 0.98 | 0.08 | 0.50 | 0.62 | 0.85 | 0.08 | 0.71 | 0.48 | 0.67 |
| **TNFRSF9** | -0.30 | -1.69 | 0.10 | 0.63 | -0.24 | -1.11 | 0.27 | 0.68 | -0.09 | -0.46 | 0.65 | 0.87 | -0.07 | -0.37 | 0.71 | 0.83 |
| **TNFSF10** | -0.12 | -1.25 | 0.22 | 0.78 | 0.07 | 0.59 | 0.56 | 0.86 | 0.01 | 0.09 | 0.93 | 0.99 | 0.02 | 0.15 | 0.88 | 0.95 |
| **TNFSF11** | 0.25 | 1.44 | 0.15 | 0.70 | 0.39 | 2.05 | 0.04 | 0.37 | 0.35 | 1.97 | 0.05 | 0.21 | 0.45 | 2.67 | 0.01 | 0.07 |
| **TNFSF12** | 0.00 | 0.01 | 0.99 | 1.00 | -0.06 | -0.44 | 0.66 | 0.90 | -0.07 | -0.57 | 0.57 | 0.83 | 0.10 | 0.91 | 0.37 | 0.60 |
| **TNFSF13** | 0.18 | 1.43 | 0.16 | 0.70 | 0.08 | 0.65 | 0.52 | 0.84 | 0.24 | 2.20 | 0.03 | 0.15 | 0.20 | 1.88 | 0.07 | 0.23 |
| **TNFSF14** | 0.03 | 0.22 | 0.83 | 0.99 | 0.09 | 0.61 | 0.54 | 0.85 | -0.05 | -0.39 | 0.70 | 0.90 | 0.11 | 0.81 | 0.42 | 0.64 |
| **TNFSF15** | 0.08 | 0.84 | 0.40 | 0.89 | 0.10 | 0.88 | 0.38 | 0.75 | -0.14 | -0.80 | 0.43 | 0.72 | 0.21 | 2.20 | 0.03 | 0.14 |
| **TNFSF18** | 0.00 | 0.04 | 0.97 | 0.99 | 0.16 | 1.54 | 0.13 | 0.50 | 0.07 | 0.80 | 0.43 | 0.72 | 0.07 | 0.89 | 0.38 | 0.60 |
| **TNFSF4** | -0.08 | -0.80 | 0.43 | 0.89 | -0.04 | -0.33 | 0.74 | 0.95 | -0.07 | -0.73 | 0.47 | 0.75 | -0.13 | -1.35 | 0.18 | 0.39 |
| **TNFSF8** | -0.02 | -0.14 | 0.89 | 0.99 | 0.19 | 1.19 | 0.24 | 0.65 | 0.16 | 1.36 | 0.18 | 0.47 | 0.08 | 0.79 | 0.43 | 0.65 |
| **TNFSF9** | -0.46 | -2.88 | 0.01 | 0.33 | -0.44 | -2.65 | 0.01 | 0.19 | -0.39 | -2.48 | 0.02 | 0.10 | -0.19 | -1.25 | 0.22 | 0.44 |
| **TREM1** | 0.14 | 1.06 | 0.29 | 0.86 | 0.22 | 1.55 | 0.13 | 0.50 | 0.15 | 1.27 | 0.21 | 0.50 | 0.27 | 2.45 | 0.02 | 0.09 |
| **TREM2** | 0.20 | 1.24 | 0.22 | 0.78 | 0.49 | 2.81 | 0.01 | 0.16 | 0.40 | 2.63 | 0.01 | 0.08 | 0.40 | 3.06 | 0.00 | 0.03 |
| **TSLP** | 0.25 | 1.89 | 0.06 | 0.56 | 0.36 | 2.43 | 0.02 | 0.24 | 0.38 | 2.34 | 0.02 | 0.13 | 0.36 | 2.96 | 0.00 | 0.04 |
| **VCAM1** | -0.01 | -0.12 | 0.91 | 0.99 | 0.07 | 0.58 | 0.57 | 0.86 | 0.10 | 1.10 | 0.27 | 0.57 | 0.14 | 1.79 | 0.08 | 0.26 |
| **VEGFA** | 0.24 | 1.54 | 0.13 | 0.70 | -0.07 | -0.43 | 0.67 | 0.90 | 0.29 | 2.03 | 0.05 | 0.20 | 0.20 | 1.34 | 0.19 | 0.39 |
| **VEGFC** | -0.04 | -0.42 | 0.68 | 0.97 | 0.01 | 0.08 | 0.94 | 0.99 | 0.07 | 0.72 | 0.48 | 0.75 | 0.14 | 1.53 | 0.13 | 0.32 |
| **VEGFD** | -0.19 | -1.49 | 0.14 | 0.70 | 0.19 | 1.20 | 0.23 | 0.65 | 0.20 | 1.28 | 0.21 | 0.49 | 0.01 | 0.11 | 0.91 | 0.95 |
| **VSNL1** | -0.01 | -0.08 | 0.93 | 0.99 | 0.29 | 2.07 | 0.04 | 0.36 | 0.15 | 1.25 | 0.22 | 0.51 | 0.16 | 1.49 | 0.14 | 0.33 |
| **VSTM1** | -0.01 | -0.02 | 0.98 | 0.99 | 0.74 | 1.85 | 0.07 | 0.43 | 0.38 | 1.07 | 0.29 | 0.59 | 0.59 | 1.64 | 0.11 | 0.30 |
| **WNT16** | 0.07 | 0.41 | 0.69 | 0.97 | 0.18 | 1.07 | 0.29 | 0.68 | 0.14 | 0.87 | 0.39 | 0.68 | 0.33 | 1.86 | 0.07 | 0.23 |
| **WNT7A** | -0.29 | -1.25 | 0.21 | 0.78 | -0.40 | -1.49 | 0.14 | 0.52 | -0.16 | -0.71 | 0.48 | 0.75 | -0.14 | -0.62 | 0.54 | 0.73 |
